# Receipt of anti-SARS-CoV-2 pharmacotherapies among non-hospitalized U.S. Veterans with COVID-19, January 2022 to January 2023

**DOI:** 10.1101/2023.05.03.23289479

**Authors:** Lei Yan, Elani Streja, Yuli Li, Nallakkandi Rajeevan, Mazhgan Rowneki, Kristin Berry, Denise M. Hynes, Francesca Cunningham, Grant D. Huang, Mihaela Aslan, George N. Ioannou, Kristina L. Bajema

## Abstract

**IMPORTANCE:** Several pharmacotherapies have been authorized to treat non-hospitalized persons with symptomatic COVID-19. Longitudinal information on their use is needed.

**OBJECTIVE:** To analyze trends and factors related to prescription of outpatient COVID-19 pharmacotherapies within the Veterans Health Administration (VHA).

**DESIGN, SETTINGS, AND PARTICIPANTS:** This cohort study evaluated non-hospitalized veterans in VHA care who tested positive for SARS-CoV-2 from January 2022 through January 2023, using VHA and linked Community Care and Medicare databases.

**EXPOSURES:** Demographic characteristics, regional and local systems of care including Veterans Integrated Services Networks (VISNs), underlying medical conditions, COVID-19 vaccination.

**MAIN OUTCOMES AND MEASURES:** Monthly receipt of any COVID-19 pharmacotherapy (nirmatrelvir-ritonavir, molnupiravir, sotrovimab, or bebtelovimab) was described. Multivariable logistic regression was used to identify factors independently associated with receipt of any versus no COVID-19 pharmacotherapy.

**RESULTS:** Among 285,710 veterans (median [IQR] age, 63.1 [49.9-73.7] years; 247,358 (86.6%) male; 28,444 (10%) Hispanic; 198,863 (72.7%) White; 61,269 (22.4%) Black) who tested positive for SARS-CoV-2 between January 2022 and January 2023, the proportion receiving any pharmacotherapy increased from 3.2% (3,285/102,343) in January 2022 to 23.9% (5,180/21,688) in August 2022, and declined slightly to 20.8% (2,194/10,551) by January 2023. Across VISNs, the range in proportion of test-positive patients who received nirmatrelvir-ritonavir or molnupiravir during January 2023 was 5.9 to 21.4% and 2.1 to 11.1%, respectively. Veterans receiving any treatment were more likely to be older (adjusted odds ratio [aOR], 1.18, 95% CI 1.14-1.22 for 65 to 74 versus 50 to 64 years; aOR 1.19, 95% CI 1.15-1.23 for 75 versus 50 to 64 years), have a higher Charlson Comorbidity Index (CCI) (aOR 1.52, 95% CI 1.44-1.59 for CCI ≥6 versus 0), and be vaccinated against COVID-19 (aOR 1.25, 95% CI 1.19-1.30 for primary versus no vaccination; aOR 1.47, 95% CI 1.42-1.53 for booster versus no vaccination). Compared with White veterans, Black veterans (aOR 1.06, 95% CI 1.02 to 1.09) were more likely to receive treatment, and compared with non-Hispanic veterans, Hispanic veterans (aOR 1.06, 95% CI 1.01-1.11) were more likely to receive treatment.

**CONCLUSIONS AND RELEVANCE:** Among veterans who tested positive for SARS-CoV-2 between January 2022 and January 2023, prescription of outpatient COVID-19 pharmacotherapies peaked in August 2022 and declined thereafter. There remain large regional differences in patterns of nirmatrelvir-ritonavir and molnupiravir use.

## Introduction

Several SARS-CoV-2 antiviral agents are recommended in the United States for the treatment of COVID-19 in non-hospitalized adults at risk for progressing to severe disease.^1^ These include ritonavir-boosted nirmatrelvir, remdesivir, and molnupiravir, which received US Food and Drug Administration (FDA) Emergency Use Authorization (EUA) between December 2021 and January 2022.^2^ Collectively, these pharmacotherapies have been demonstrated to aid clinical recovery and reduce the risk of hospitalization and death.^3–7^ Following slow early uptake, by April 2023, 8.9 million courses of nirmatrelvir-ritonavir and 1.3 million courses of molnupiravir had been administered across the US.^8–11^ Despite this, challenges that impede broader uptake among vulnerable populations remain, and information on use more than one year after authorization remains limited.

The Veterans Health Administration (VHA), operated by the US Department of Veterans Affairs (VA), is the largest integrated healthcare system in the US and serves more than 9 million enrolled veterans each year at 171 medical centers and 1,113 outpatient sites of care.^12^ COVID-19 pharmacotherapies under EUA are allocated across VHA pharmacies through a national distribution system coordinated by the Pharmacy Benefits Management Services (PBM). This system provides an opportunity to examine how these therapies have been allocated to patients infected with SARS-CoV-2 over time.^13^ We sought to describe trends and factors associated with prescription of COVID-19 pharmacotherapies from January 2022 through January 2023, focusing on nirmatrelvir-ritonavir, molnupiravir, and historically available neutralizing monoclonal antibodies prescribed during this period.

## Methods

### Data Sources

We used VA’s COVID-19 Shared Data Resource (CSDR), provisioned by the VA Informatics and Computing Infrastructure (VINCI), which integrates multiple data sources to provide patient-level COVID-19-related information on VA enrollees.^14^ Positive SARS-CoV-2 tests among VA enrollees are identified by the VA National Surveillance Tool (NST) and provided to CSDR to support national VA research and operations needs.^15^ NST identifies VA enrollees with a laboratory-confirmed, positive SARS-CoV-2 nucleic acid amplification or antigen test performed within VHA or who had evidence of testing positive outside of VHA and documented in VHA clinical records (either recorded in templated notes yielding structured data or by natural language processing of the electronic health record (EHR) which is confirmed by manual chart review).

COVID-19 pharmacotherapies were ascertained using three sources: (i) the VA Corporate Data Warehouse (CDW), a linked database of the VHA EHR that contains prescription records from inpatient and outpatient care, (ii) Medicare claims data from the Centers for Medicare and Medicaid Services (CMS), provided by the VA Information Resource Center (VIReC), which includes claims for veterans who utilized Medicare, and (iii) COVID-19 monoclonal antibody claims data from the VA Community Care program, which coordinates and reimburses local care provided outside VHA. For this analysis, both CMS Medicare and VA Community Care data were available through September 30, 2022. We also used the CDW to obtain patient-level demographic, clinical, and administrative data. Hospitalization data were obtained from CDW as well as CMS Medicare. The study was approved by the VA Central Institutional Review Board and followed the Strengthening the Reporting of Observational Studies in Epidemiology (STROBE) reporting guideline.

### Study Population and Baseline Characteristics

We identified veterans aged 18 years or older with a first positive SARS-CoV-2 test between January 1, 2022, and January 31, 2023. The study population was limited to VA enrollees who had at least one VHA primary care outpatient encounter during the 18 months before the positive SARS-CoV-2 test and were not hospitalized on or within 7 days prior to the positive test.

Using the date of the positive SARS-CoV-2 test as the index date, we ascertained baseline demographic characteristics including race and ethnicity (associated with COVID-19 care) as reported in the VA EHR, VHA facility and Veterans Integrated Service Network (VISN, the 18 regional systems of care)^16^ associated with the SARS-CoV-2 test, and rurality of residence based on the Rural-Urban Commuting Areas (RUCA) system.^17^ We also determined smoking status, alcohol or substance use disorder, underlying medical conditions including immunocompromised status, Charlson comorbidity index (CCI), and COVID-19 vaccination status as previously described.^8,18^

### COVID-19 Pharmacotherapies

We identified receipt nirmatrelvir-ritonavir, molnupiravir, and two anti-SARS-CoV-2 monoclonal antibodies in use during the study period (sotrovimab and bebtelovimab, designated as the monoclonal antibody group), as captured by VHA CDW prescriptions, CMS-Medicare, and VA Community Care claims. Due to circulation of Omicron variants with reduced sensitivity to monoclonal antibodies, FDA authorization for sotrovimab was removed in April 2022, and authorization for bebtelovimab was removed in November 2022.^2^ By January 2022, use of both bamlanivimab and etesevimab as well as casirivimab and imdevimab was already very limited due to reduced activity against Omicron variants; we therefore did not include these in the monoclonal antibody group.^19^ Although remdesivir was authorized by the FDA for the treatment of COVID-19 in non-hospitalized patients in January 2022,^20^ the number of individuals who received outpatient remdesivir was small and not readily distinguished from inpatient remdesivir, thus we did not include this as a separate treatment group. Eligible veterans were assigned to a treatment group based on the first treatment received within ±7 days of their first positive SARS-CoV-2 test. Individuals who did not receive any outpatient COVID-19 pharmacotherapy within ±7 days of their test were assigned to the no treatment group.

### Statistical Analyses

Among test-positive patients, we calculated the proportion prescribed each COVID-19 pharmacotherapy between January 2022 and January 2023 by month and according to VISN, VA facility, demographic and clinical characteristics. To investigate factors associated with receipt of any treatment versus no treatment, we estimated unadjusted and adjusted odds ratios (aORs) with 95% confidence intervals (CIs) with binomial logistic regression models using data from April 2022 to January 2023, when the relative proportion of test-positive veterans receiving outpatient COVID-19 pharmacotherapies had stabilized. Models were adjusted for age, sex, race, ethnicity, VISN, and CCI. To avoid overadjustment, we did not include CCI when evaluating individual underlying conditions. Final models utilized complete data for all included covariates. Analyses were conducted using R version 4.1.2.

## Results

### Patient Characteristics

Between January 2022 and January 2023, 285,710 VA enrollees (median [IQR] age 63.1 [49.9-73.7]; 247,358 (86.6%) male; 28,444 (10%) Hispanic; 198,863 (72.7%) White; 61,269 (22.4%) Black) tested positive for SARS-CoV-2 who fulfilled study inclusion criteria (**Figure 1, Table 1**). During January 2022, more than 102,343 veterans tested positive for SARS-CoV-2; this decreased to 20,450 the following month and remained relatively stable thereafter (**Figure 2**). Of all test-positive persons, 26,677 (9.3%) received nirmatrelvir-ritonavir, 9,003 (3.2%) received molnupiravir, 4,784 (1.7%) received monoclonal antibodies (sotrovimab or bebtelovimab), and 239,563 (83.8%) did not receive treatment. The remaining 5,683 (2.0%) received other COVID-19 pharmacotherapies (5,058 [1.8%] remdesivir and 638 [0.3%] other monoclonal antibodies) and were not included in any treatment or no treatment groups. Most pharmacotherapies were identified from VA CDW (96% of all nirmatrelvir-ritonavir prescriptions, 96% of all molnupiravir prescriptions, and 79% of all monoclonal antibody treatments, **Supplemental Table 1, Supplemental Figure 1**). CMS Medicare data contributed to 4% of all nirmatrelvir-ritonavir and molnupiravir prescriptions and 9% of monoclonal antibody treatments. The remaining 11% of monoclonal antibody treatments were from VA Community Care data.

**Figure 1.**
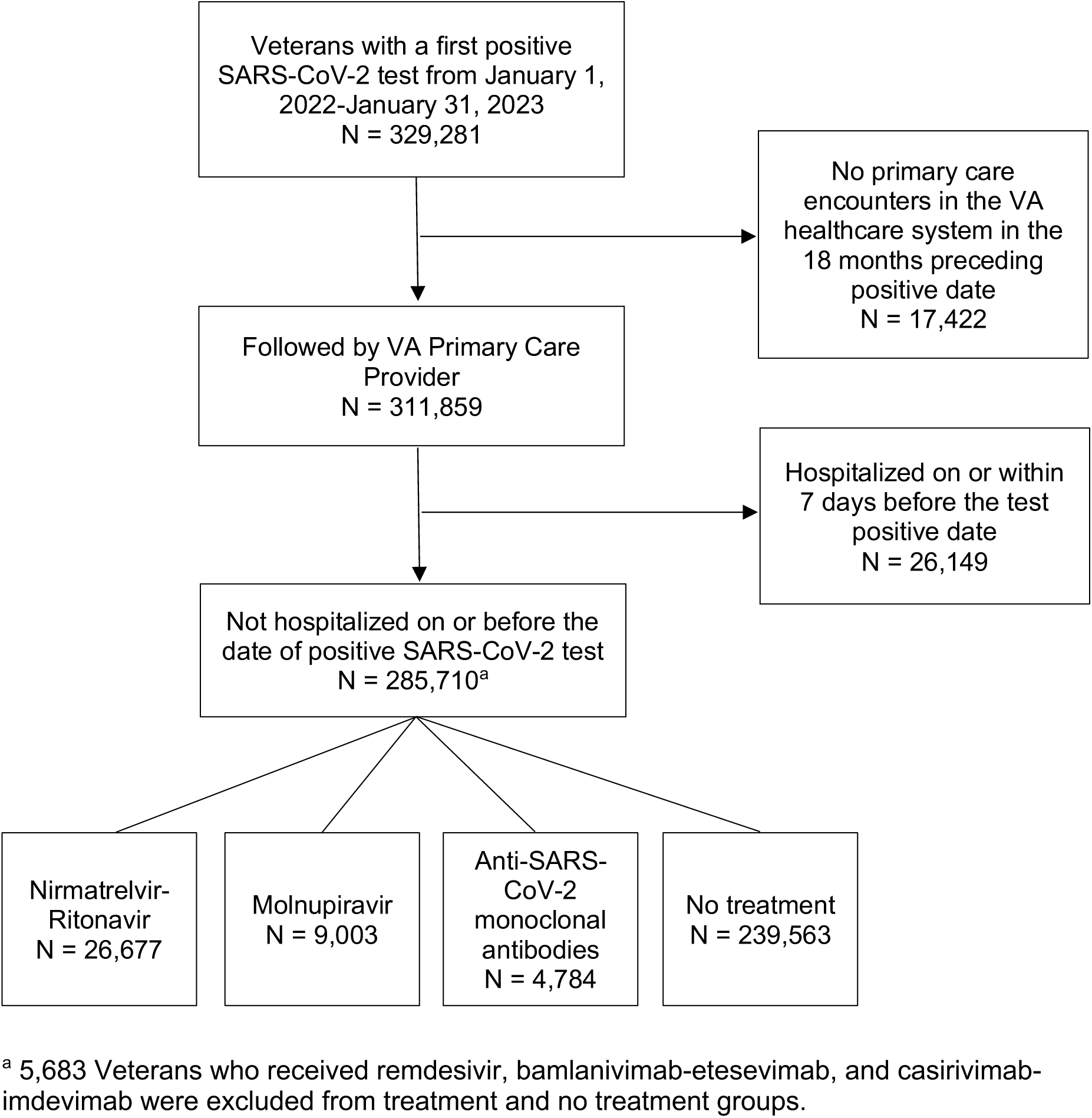
VA enrollees with a first positive SARS-CoV-2 test from January 1, 2022 to January 31, 2023 included in the study ^a^ 5,683 Veterans who received remdesivir, bamlanivimab-etesevimab, and casirivimab-imdevimab were excluded from treatment and no treatment groups.

**Figure 2.**
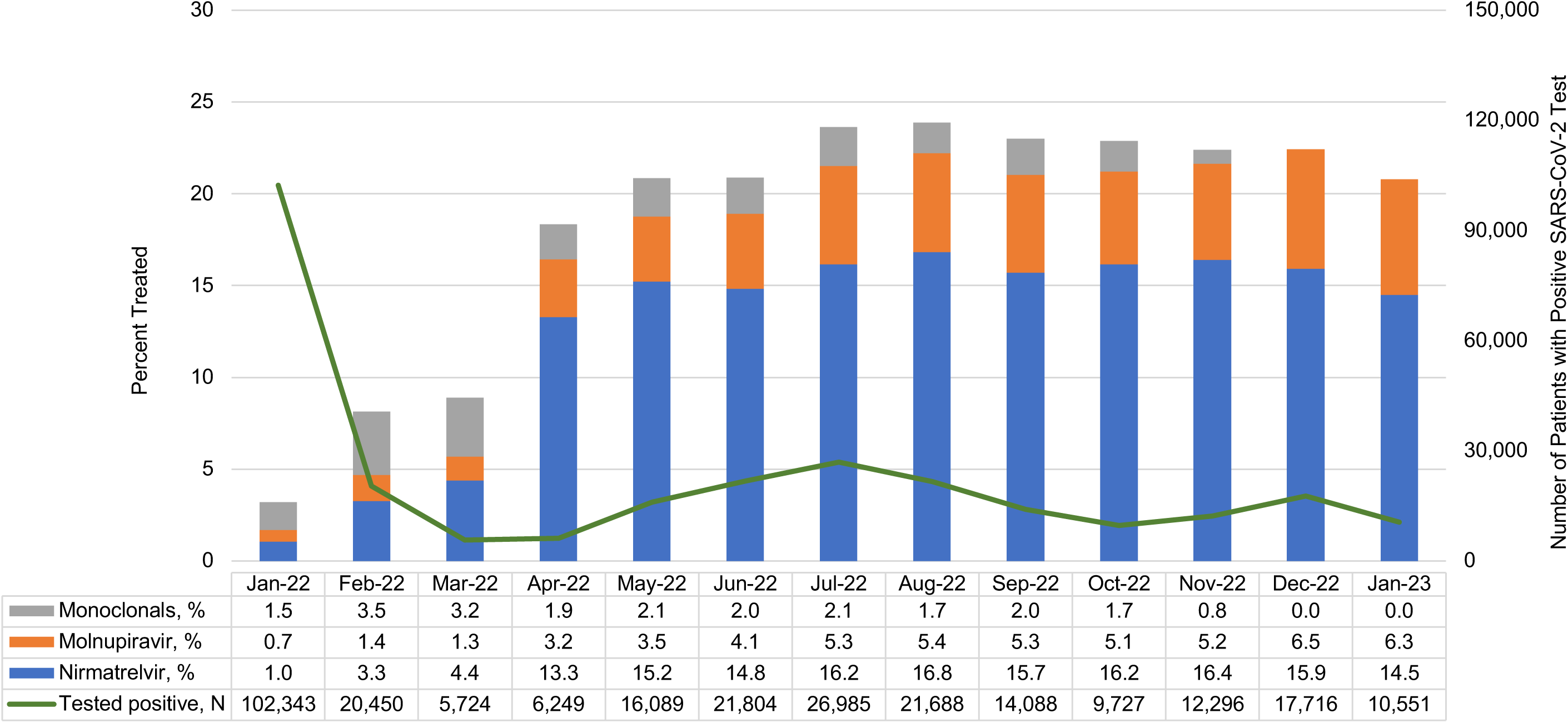
Distribution of COVID-19 pharmacotherapies administered among SARS-CoV-2 test-positive patients in the Veterans Health Administration, January 2022 to January 2023.

**Table 1.**
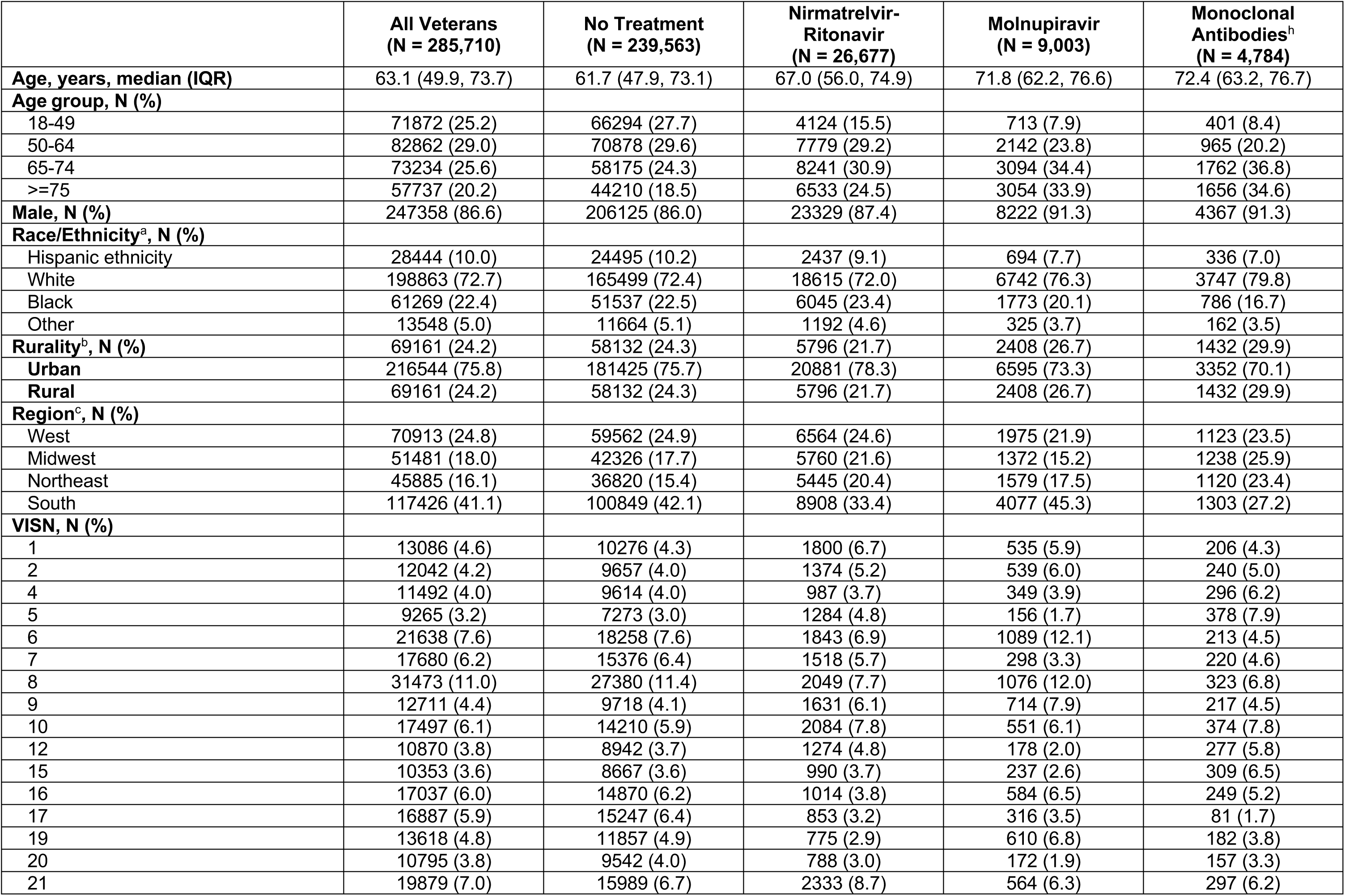
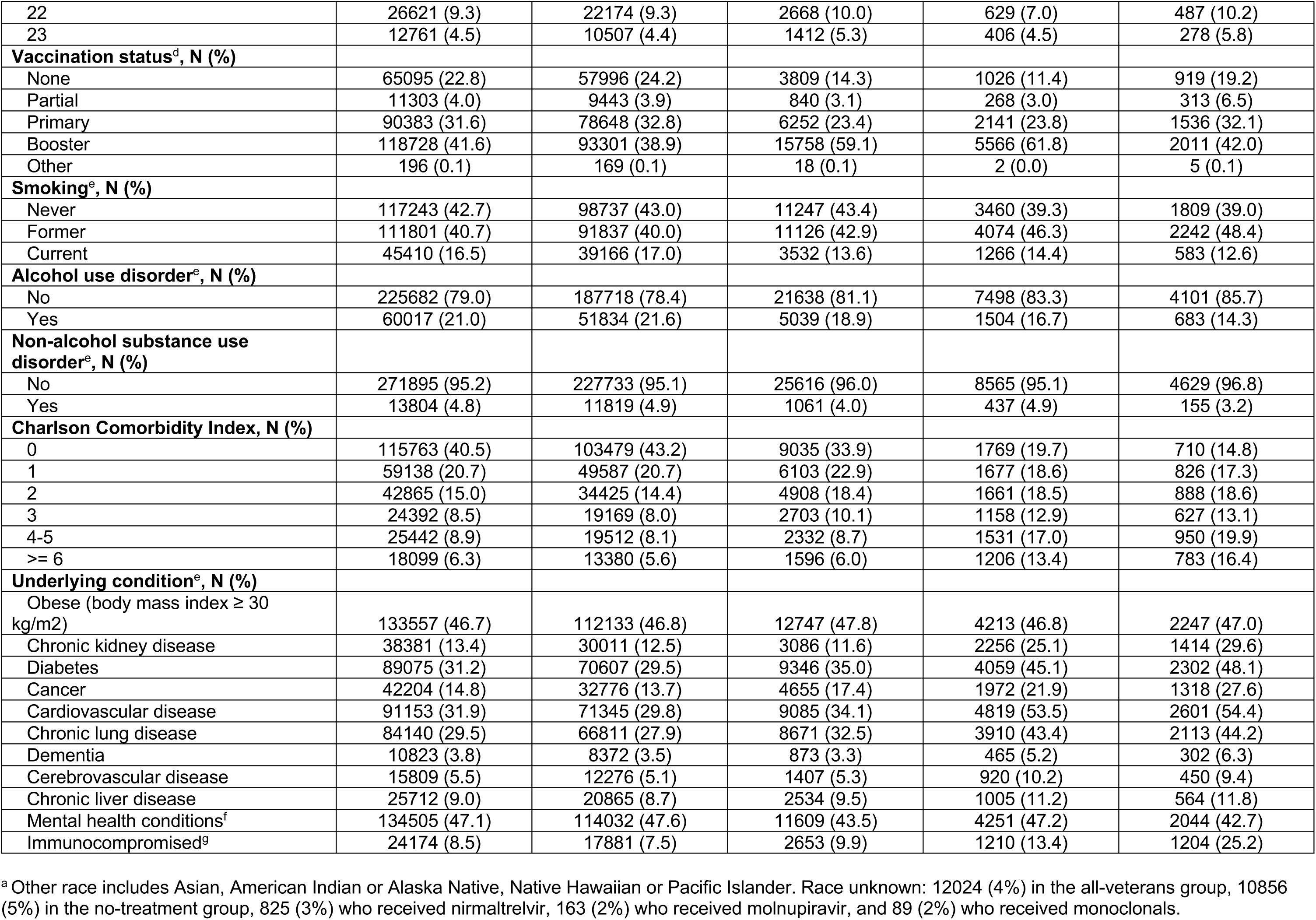
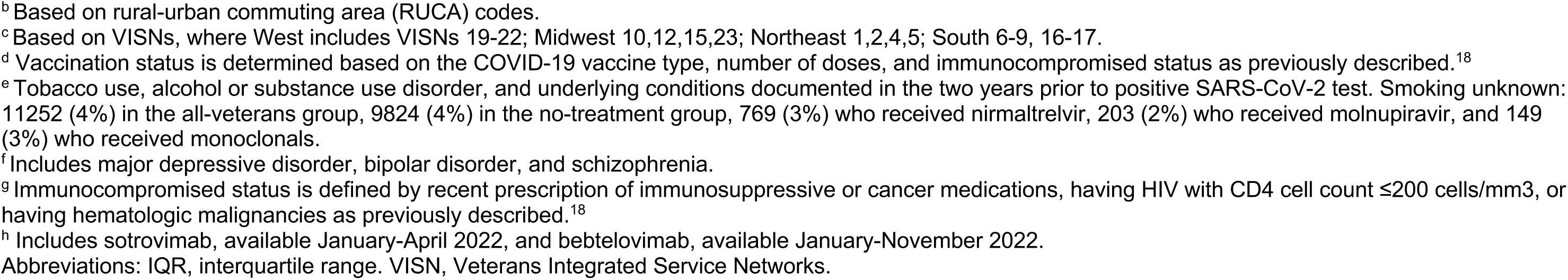
Characteristics of Veterans testing positive for SARS-CoV-2 by receipt of COVID-19 pharmacotherapy, January 2022 to January 2023

|  | **All Veterans**<br>**(N = 285,710)** | **No Treatment**<br>**(N = 239,563)** | **Nirmatrelvir-**<br>**Ritonavir**<br>**(N = 26,677)** | **Molnupiravir**<br>**(N = 9,003)** | **Monoclonal**<br>**Antibodies<sup>h</sup>**<br>**(N = 4,784)** |
| --- | --- | --- | --- | --- | --- |
| **Age, years, median (IQR)** | 63.1 (49.9, 73.7) | 61.7 (47.9, 73.1) | 67.0 (56.0, 74.9) | 71.8 (62.2, 76.6) | 72.4 (63.2, 76.7) |
| **Age group, N (%)** |  |  |  |  |  |
| 18-49 | 71872 (25.2) | 66294 (27.7) | 4124 (15.5) | 713 (7.9) | 401 (8.4) |
| 50-64 | 82862 (29.0) | 70878 (29.6) | 7779 (29.2) | 2142 (23.8) | 965 (20.2) |
| 65-74 | 73234 (25.6) | 58175 (24.3) | 8241 (30.9) | 3094 (34.4) | 1762 (36.8) |
| >=75 | 57737 (20.2) | 44210 (18.5) | 6533 (24.5) | 3054 (33.9) | 1656 (34.6) |
| **Male, N (%)** | 247358 (86.6) | 206125 (86.0) | 23329 (87.4) | 8222 (91.3) | 4367 (91.3) |
| **Race/Ethnicity<sup>a</sup>, N (%)** |  |  |  |  |  |
| Hispanic ethnicity | 28444 (10.0) | 24495 (10.2) | 2437 (9.1) | 694 (7.7) | 336 (7.0) |
| White | 198863 (72.7) | 165499 (72.4) | 18615 (72.0) | 6742 (76.3) | 3747 (79.8) |
| Black | 61269 (22.4) | 51537 (22.5) | 6045 (23.4) | 1773 (20.1) | 786 (16.7) |
| Other | 13548 (5.0) | 11664 (5.1) | 1192 (4.6) | 325 (3.7) | 162 (3.5) |
| **Rurality<sup>b</sup>, N (%)** | 69161 (24.2) | 58132 (24.3) | 5796 (21.7) | 2408 (26.7) | 1432 (29.9) |
| **Urban** | 216544 (75.8) | 181425 (75.7) | 20881 (78.3) | 6595 (73.3) | 3352 (70.1) |
| **Rural** | 69161 (24.2) | 58132 (24.3) | 5796 (21.7) | 2408 (26.7) | 1432 (29.9) |
| **Region<sup>c</sup>, N (%)** |  |  |  |  |  |
| West | 70913 (24.8) | 59562 (24.9) | 6564 (24.6) | 1975 (21.9) | 1123 (23.5) |
| Midwest | 51481 (18.0) | 42326 (17.7) | 5760 (21.6) | 1372 (15.2) | 1238 (25.9) |
| Northeast | 45885 (16.1) | 36820 (15.4) | 5445 (20.4) | 1579 (17.5) | 1120 (23.4) |
| South | 117426 (41.1) | 100849 (42.1) | 8908 (33.4) | 4077 (45.3) | 1303 (27.2) |
| **VISN, N (%)** |  |  |  |  |  |
| 1 | 13086 (4.6) | 10276 (4.3) | 1800 (6.7) | 535 (5.9) | 206 (4.3) |
| 2 | 12042 (4.2) | 9657 (4.0) | 1374 (5.2) | 539 (6.0) | 240 (5.0) |
| 4 | 11492 (4.0) | 9614 (4.0) | 987 (3.7) | 349 (3.9) | 296 (6.2) |
| 5 | 9265 (3.2) | 7273 (3.0) | 1284 (4.8) | 156 (1.7) | 378 (7.9) |
| 6 | 21638 (7.6) | 18258 (7.6) | 1843 (6.9) | 1089 (12.1) | 213 (4.5) |
| 7 | 17680 (6.2) | 15376 (6.4) | 1518 (5.7) | 298 (3.3) | 220 (4.6) |
| 8 | 31473 (11.0) | 27380 (11.4) | 2049 (7.7) | 1076 (12.0) | 323 (6.8) |
| 9 | 12711 (4.4) | 9718 (4.1) | 1631 (6.1) | 714 (7.9) | 217 (4.5) |
| 10 | 17497 (6.1) | 14210 (5.9) | 2084 (7.8) | 551 (6.1) | 374 (7.8) |
| 12 | 10870 (3.8) | 8942 (3.7) | 1274 (4.8) | 178 (2.0) | 277 (5.8) |
| 15 | 10353 (3.6) | 8667 (3.6) | 990 (3.7) | 237 (2.6) | 309 (6.5) |
| 16 | 17037 (6.0) | 14870 (6.2) | 1014 (3.8) | 584 (6.5) | 249 (5.2) |
| 17 | 16887 (5.9) | 15247 (6.4) | 853 (3.2) | 316 (3.5) | 81 (1.7) |
| 19 | 13618 (4.8) | 11857 (4.9) | 775 (2.9) | 610 (6.8) | 182 (3.8) |
| 20 | 10795 (3.8) | 9542 (4.0) | 788 (3.0) | 172 (1.9) | 157 (3.3) |
| 21 | 19879 (7.0) | 15989 (6.7) | 2333 (8.7) | 564 (6.3) | 297 (6.2) |
| 22 | 26621 (9.3) | 22174 (9.3) | 2668 (10.0) | 629 (7.0) | 487 (10.2) |
| 23 | 12761 (4.5) | 10507 (4.4) | 1412 (5.3) | 406 (4.5) | 278 (5.8) |
| **Vaccination status<sup>d</sup>, N (%)** |  |  |  |  |  |
| None | 65095 (22.8) | 57996 (24.2) | 3809 (14.3) | 1026 (11.4) | 919 (19.2) |
| Partial | 11303 (4.0) | 9443 (3.9) | 840 (3.1) | 268 (3.0) | 313 (6.5) |
| Primary | 90383 (31.6) | 78648 (32.8) | 6252 (23.4) | 2141 (23.8) | 1536 (32.1) |
| Booster | 118728 (41.6) | 93301 (38.9) | 15758 (59.1) | 5566 (61.8) | 2011 (42.0) |
| Other | 196 (0.1) | 169 (0.1) | 18 (0.1) | 2 (0.0) | 5 (0.1) |
| **Smoking<sup>e</sup>, N (%)** |  |  |  |  |  |
| Never | 117243 (42.7) | 98737 (43.0) | 11247 (43.4) | 3460 (39.3) | 1809 (39.0) |
| Former | 111801 (40.7) | 91837 (40.0) | 11126 (42.9) | 4074 (46.3) | 2242 (48.4) |
| Current | 45410 (16.5) | 39166 (17.0) | 3532 (13.6) | 1266 (14.4) | 583 (12.6) |
| **Alcohol use disorder<sup>e</sup>, N (%)** |  |  |  |  |  |
| No | 225682 (79.0) | 187718 (78.4) | 21638 (81.1) | 7498 (83.3) | 4101 (85.7) |
| Yes | 60017 (21.0) | 51834 (21.6) | 5039 (18.9) | 1504 (16.7) | 683 (14.3) |
| **Non-alcohol substance use disorder<sup>e</sup>, N (%)** |  |  |  |  |  |
| No | 271895 (95.2) | 227733 (95.1) | 25616 (96.0) | 8565 (95.1) | 4629 (96.8) |
| Yes | 13804 (4.8) | 11819 (4.9) | 1061 (4.0) | 437 (4.9) | 155 (3.2) |
| **Charlson Comorbidity Index, N (%)** |  |  |  |  |  |
| 0 | 115763 (40.5) | 103479 (43.2) | 9035 (33.9) | 1769 (19.7) | 710 (14.8) |
| 1 | 59138 (20.7) | 49587 (20.7) | 6103 (22.9) | 1677 (18.6) | 826 (17.3) |
| 2 | 42865 (15.0) | 34425 (14.4) | 4908 (18.4) | 1661 (18.5) | 888 (18.6) |
| 3 | 24392 (8.5) | 19169 (8.0) | 2703 (10.1) | 1158 (12.9) | 627 (13.1) |
| 4-5 | 25442 (8.9) | 19512 (8.1) | 2332 (8.7) | 1531 (17.0) | 950 (19.9) |
| >= 6 | 18099 (6.3) | 13380 (5.6) | 1596 (6.0) | 1206 (13.4) | 783 (16.4) |
| **Underlying condition<sup>e</sup>, N (%)** |  |  |  |  |  |
| Obese (body mass index ≥ 30 kg/m<sup>2</sup>) | 133557 (46.7) | 112133 (46.8) | 12747 (47.8) | 4213 (46.8) | 2247 (47.0) |
| Chronic kidney disease | 38381 (13.4) | 30011 (12.5) | 3086 (11.6) | 2256 (25.1) | 1414 (29.6) |
| Diabetes | 89075 (31.2) | 70607 (29.5) | 9346 (35.0) | 4059 (45.1) | 2302 (48.1) |
| Cancer | 42204 (14.8) | 32776 (13.7) | 4655 (17.4) | 1972 (21.9) | 1318 (27.6) |
| Cardiovascular disease | 91153 (31.9) | 71345 (29.8) | 9085 (34.1) | 4819 (53.5) | 2601 (54.4) |
| Chronic lung disease | 84140 (29.5) | 66811 (27.9) | 8671 (32.5) | 3910 (43.4) | 2113 (44.2) |
| Dementia | 10823 (3.8) | 8372 (3.5) | 873 (3.3) | 465 (5.2) | 302 (6.3) |
| Cerebrovascular disease | 15809 (5.5) | 12276 (5.1) | 1407 (5.3) | 920 (10.2) | 450 (9.4) |
| Chronic liver disease | 25712 (9.0) | 20865 (8.7) | 2534 (9.5) | 1005 (11.2) | 564 (11.8) |
| Mental health conditions<sup>f</sup> | 134505 (47.1) | 114032 (47.6) | 11609 (43.5) | 4251 (47.2) | 2044 (42.7) |
| Immunocompromised<sup>g</sup> | 24174 (8.5) | 17881 (7.5) | 2653 (9.9) | 1210 (13.4) | 1204 (25.2) |

<sup>a</sup> Other race includes Asian, American Indian or Alaska Native, Native Hawaiian or Pacific Islander. Race unknown: 12024 (4%) in the all-veterans group, 10856 (5%) in the no-treatment group, 825 (3%) who received nirmaltrelvir, 163 (2%) who received molnupiravir, and 89 (2%) who received monoclonals.
<sup>b</sup> Based on rural-urban commuting area (RUCA) codes.
<sup>c</sup> Based on VISNs, where West includes VISNs 19-22; Midwest 10,12,15,23; Northeast 1,2,4,5; South 6-9, 16-17.
<sup>d</sup> Vaccination status is determined based on the COVID-19 vaccine type, number of doses, and immunocompromised status as previously described.<sup>18</sup>
<sup>e</sup> Tobacco use, alcohol or substance use disorder, and underlying conditions documented in the two years prior to positive SARS-CoV-2 test. Smoking unknown: 11252 (4%) in the all-veterans group, 9824 (4%) in the no-treatment group, 769 (3%) who received nirmaltrelvir, 203 (2%) who received molnupiravir, and 149 (3%) who received monoclonals.
<sup>f</sup> Includes major depressive disorder, bipolar disorder, and schizophrenia.
<sup>g</sup> Immunocompromised status is defined by recent prescription of immunosuppressive or cancer medications, having HIV with CD4 cell count $\leq 200$ cells/mm<sup>3</sup>, or having hematologic malignancies as previously described.<sup>18</sup>
<sup>h</sup> Includes sotrovimab, available January-April 2022, and bebtelovimab, available January-November 2022.
Abbreviations: IQR, interquartile range. VISN, Veterans Integrated Service Networks.

**Table 2.**
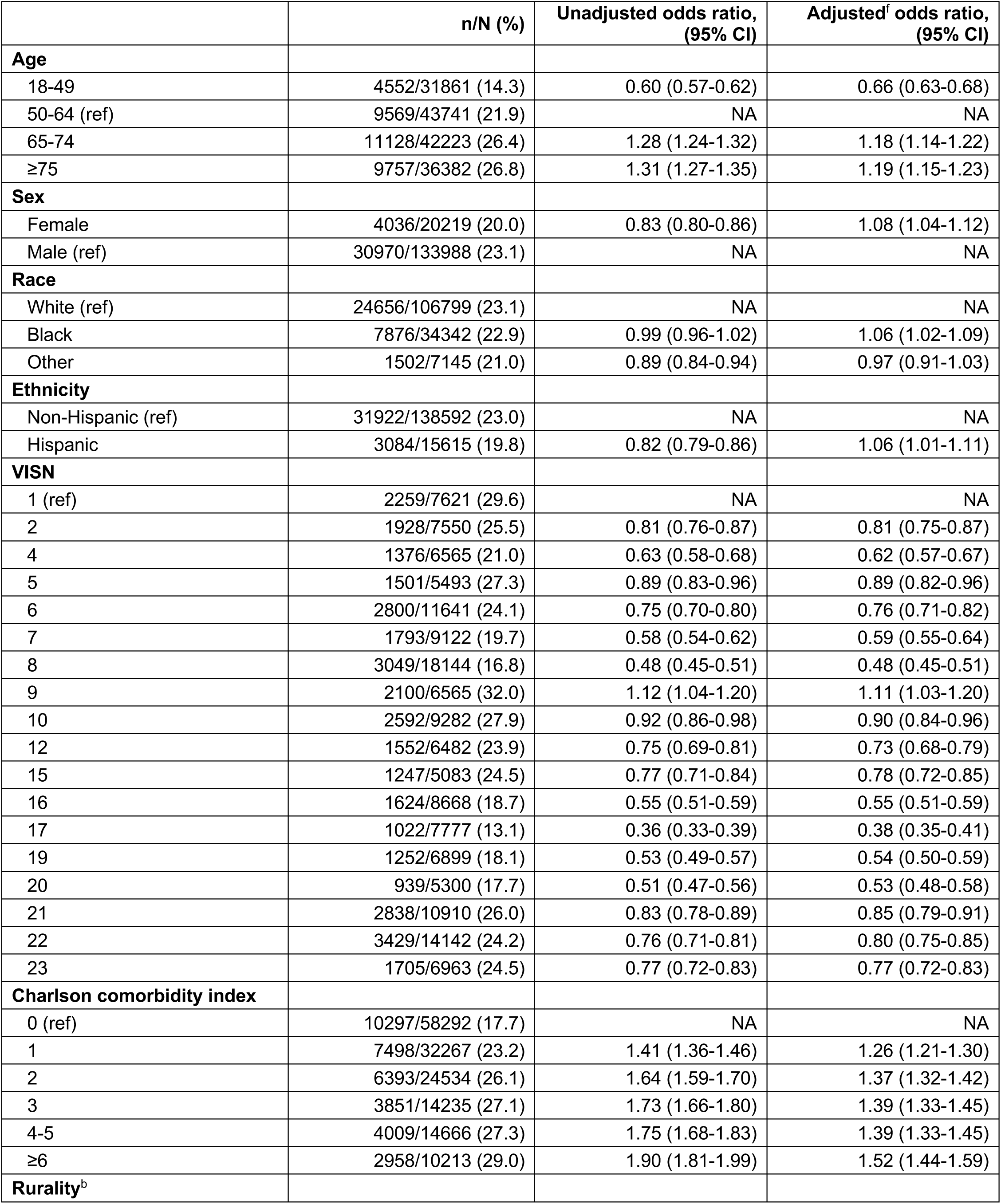
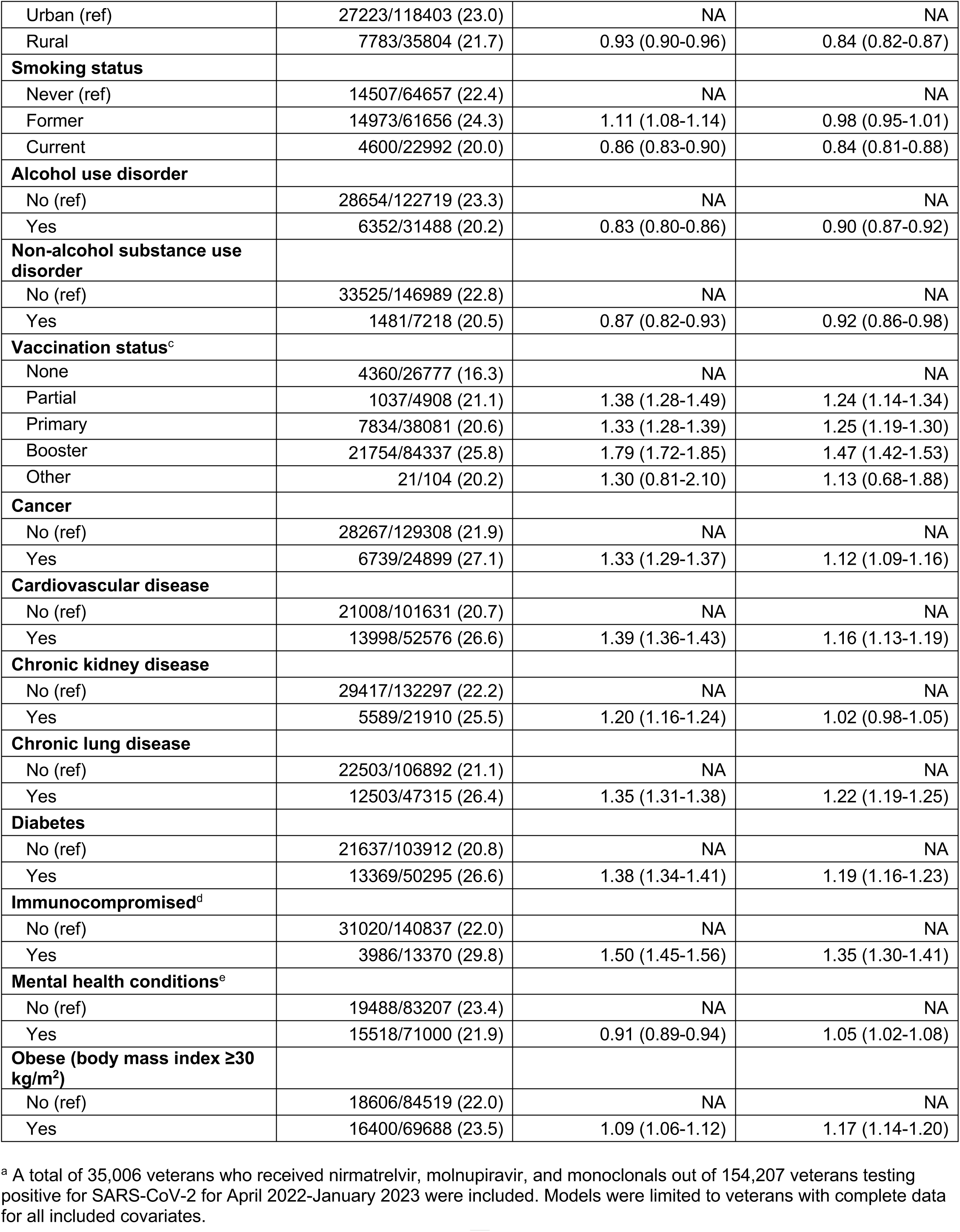
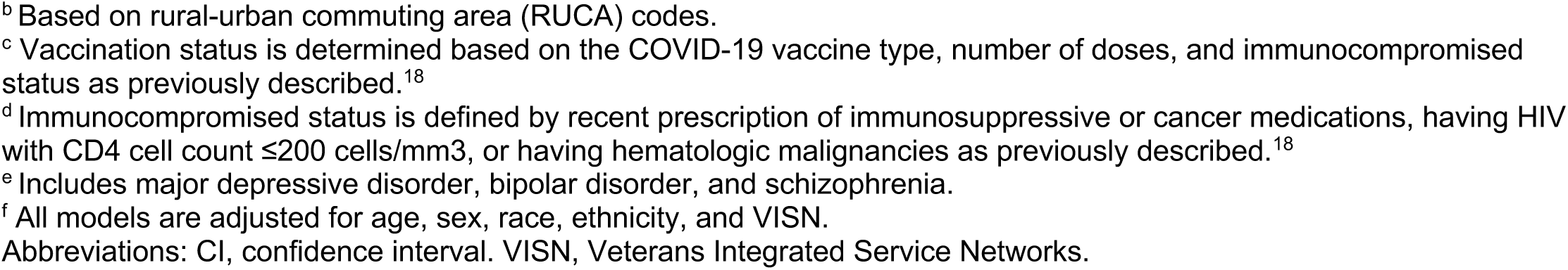
Factors associated with receipt of any COVID-19 pharmacotherapy among non-hospitalized Veterans, April 2022 to January 2023^a^

|  | n/N (%) | Unadjusted odds ratio,<br>(95% CI) | Adjusted<sup>f</sup> odds ratio,<br>(95% CI) |
| --- | --- | --- | --- |
| **Age** |  |  |  |
| 18-49 | 4552/31861 (14.3) | 0.60 (0.57-0.62) | 0.66 (0.63-0.68) |
| 50-64 (ref) | 9569/43741 (21.9) | NA | NA |
| 65-74 | 11128/42223 (26.4) | 1.28 (1.24-1.32) | 1.18 (1.14-1.22) |
| ≥75 | 9757/36382 (26.8) | 1.31 (1.27-1.35) | 1.19 (1.15-1.23) |
| **Sex** |  |  |  |
| Female | 4036/20219 (20.0) | 0.83 (0.80-0.86) | 1.08 (1.04-1.12) |
| Male (ref) | 30970/133988 (23.1) | NA | NA |
| **Race** |  |  |  |
| White (ref) | 24656/106799 (23.1) | NA | NA |
| Black | 7876/34342 (22.9) | 0.99 (0.96-1.02) | 1.06 (1.02-1.09) |
| Other | 1502/7145 (21.0) | 0.89 (0.84-0.94) | 0.97 (0.91-1.03) |
| **Ethnicity** |  |  |  |
| Non-Hispanic (ref) | 31922/138592 (23.0) | NA | NA |
| Hispanic | 3084/15615 (19.8) | 0.82 (0.79-0.86) | 1.06 (1.01-1.11) |
| **VISN** |  |  |  |
| 1 (ref) | 2259/7621 (29.6) | NA | NA |
| 2 | 1928/7550 (25.5) | 0.81 (0.76-0.87) | 0.81 (0.75-0.87) |
| 4 | 1376/6565 (21.0) | 0.63 (0.58-0.68) | 0.62 (0.57-0.67) |
| 5 | 1501/5493 (27.3) | 0.89 (0.83-0.96) | 0.89 (0.82-0.96) |
| 6 | 2800/11641 (24.1) | 0.75 (0.70-0.80) | 0.76 (0.71-0.82) |
| 7 | 1793/9122 (19.7) | 0.58 (0.54-0.62) | 0.59 (0.55-0.64) |
| 8 | 3049/18144 (16.8) | 0.48 (0.45-0.51) | 0.48 (0.45-0.51) |
| 9 | 2100/6565 (32.0) | 1.12 (1.04-1.20) | 1.11 (1.03-1.20) |
| 10 | 2592/9282 (27.9) | 0.92 (0.86-0.98) | 0.90 (0.84-0.96) |
| 12 | 1552/6482 (23.9) | 0.75 (0.69-0.81) | 0.73 (0.68-0.79) |
| 15 | 1247/5083 (24.5) | 0.77 (0.71-0.84) | 0.78 (0.72-0.85) |
| 16 | 1624/8668 (18.7) | 0.55 (0.51-0.59) | 0.55 (0.51-0.59) |
| 17 | 1022/7777 (13.1) | 0.36 (0.33-0.39) | 0.38 (0.35-0.41) |
| 19 | 1252/6899 (18.1) | 0.53 (0.49-0.57) | 0.54 (0.50-0.59) |
| 20 | 939/5300 (17.7) | 0.51 (0.47-0.56) | 0.53 (0.48-0.58) |
| 21 | 2838/10910 (26.0) | 0.83 (0.78-0.89) | 0.85 (0.79-0.91) |
| 22 | 3429/14142 (24.2) | 0.76 (0.71-0.81) | 0.80 (0.75-0.85) |
| 23 | 1705/6963 (24.5) | 0.77 (0.72-0.83) | 0.77 (0.72-0.83) |
| **Charlson comorbidity index** |  |  |  |
| 0 (ref) | 10297/58292 (17.7) | NA | NA |
| 1 | 7498/32267 (23.2) | 1.41 (1.36-1.46) | 1.26 (1.21-1.30) |
| 2 | 6393/24534 (26.1) | 1.64 (1.59-1.70) | 1.37 (1.32-1.42) |
| 3 | 3851/14235 (27.1) | 1.73 (1.66-1.80) | 1.39 (1.33-1.45) |
| 4-5 | 4009/14666 (27.3) | 1.75 (1.68-1.83) | 1.39 (1.33-1.45) |
| ≥6 | 2958/10213 (29.0) | 1.90 (1.81-1.99) | 1.52 (1.44-1.59) |
| **Rurality<sup>b</sup>** |  |  |  |
| Urban (ref) | 27223/118403 (23.0) | NA | NA |
| Rural | 7783/35804 (21.7) | 0.93 (0.90-0.96) | 0.84 (0.82-0.87) |
| **Smoking status** |  |  |  |
| Never (ref) | 14507/64657 (22.4) | NA | NA |
| Former | 14973/61656 (24.3) | 1.11 (1.08-1.14) | 0.98 (0.95-1.01) |
| Current | 4600/22992 (20.0) | 0.86 (0.83-0.90) | 0.84 (0.81-0.88) |
| **Alcohol use disorder** |  |  |  |
| No (ref) | 28654/122719 (23.3) | NA | NA |
| Yes | 6352/31488 (20.2) | 0.83 (0.80-0.86) | 0.90 (0.87-0.92) |
| **Non-alcohol substance use disorder** |  |  |  |
| No (ref) | 33525/146989 (22.8) | NA | NA |
| Yes | 1481/7218 (20.5) | 0.87 (0.82-0.93) | 0.92 (0.86-0.98) |
| **Vaccination status<sup>c</sup>** |  |  |  |
| None | 4360/26777 (16.3) | NA | NA |
| Partial | 1037/4908 (21.1) | 1.38 (1.28-1.49) | 1.24 (1.14-1.34) |
| Primary | 7834/38081 (20.6) | 1.33 (1.28-1.39) | 1.25 (1.19-1.30) |
| Booster | 21754/84337 (25.8) | 1.79 (1.72-1.85) | 1.47 (1.42-1.53) |
| Other | 21/104 (20.2) | 1.30 (0.81-2.10) | 1.13 (0.68-1.88) |
| **Cancer** |  |  |  |
| No (ref) | 28267/129308 (21.9) | NA | NA |
| Yes | 6739/24899 (27.1) | 1.33 (1.29-1.37) | 1.12 (1.09-1.16) |
| **Cardiovascular disease** |  |  |  |
| No (ref) | 21008/101631 (20.7) | NA | NA |
| Yes | 13998/52576 (26.6) | 1.39 (1.36-1.43) | 1.16 (1.13-1.19) |
| **Chronic kidney disease** |  |  |  |
| No (ref) | 29417/132297 (22.2) | NA | NA |
| Yes | 5589/21910 (25.5) | 1.20 (1.16-1.24) | 1.02 (0.98-1.05) |
| **Chronic lung disease** |  |  |  |
| No (ref) | 22503/106892 (21.1) | NA | NA |
| Yes | 12503/47315 (26.4) | 1.35 (1.31-1.38) | 1.22 (1.19-1.25) |
| **Diabetes** |  |  |  |
| No (ref) | 21637/103912 (20.8) | NA | NA |
| Yes | 13369/50295 (26.6) | 1.38 (1.34-1.41) | 1.19 (1.16-1.23) |
| **Immunocompromised<sup>d</sup>** |  |  |  |
| No (ref) | 31020/140837 (22.0) | NA | NA |
| Yes | 3986/13370 (29.8) | 1.50 (1.45-1.56) | 1.35 (1.30-1.41) |
| **Mental health conditions<sup>e</sup>** |  |  |  |
| No (ref) | 19488/83207 (23.4) | NA | NA |
| Yes | 15518/71000 (21.9) | 0.91 (0.89-0.94) | 1.05 (1.02-1.08) |
| **Obese (body mass index $\geq 30$ kg/m<sup>2</sup>)** |  |  |  |
| No (ref) | 18606/84519 (22.0) | NA | NA |
| Yes | 16400/69688 (23.5) | 1.09 (1.06-1.12) | 1.17 (1.14-1.20) |

<sup>a</sup> A total of 35,006 veterans who received nirmatrelvir, molnupiravir, and monoclonals out of 154,207 veterans testing positive for SARS-CoV-2 for April 2022-January 2023 were included. Models were limited to veterans with complete data for all included covariates.
<sup>b</sup> Based on rural-urban commuting area (RUCA) codes.
<sup>c</sup> Vaccination status is determined based on the COVID-19 vaccine type, number of doses, and immunocompromised status as previously described.<sup>18</sup>
<sup>d</sup> Immunocompromised status is defined by recent prescription of immunosuppressive or cancer medications, having HIV with CD4 cell count $\leq 200$ cells/mm<sup>3</sup>, or having hematologic malignancies as previously described.<sup>18</sup>
<sup>e</sup> Includes major depressive disorder, bipolar disorder, and schizophrenia.
<sup>f</sup> All models are adjusted for age, sex, race, ethnicity, and VISN.
Abbreviations: CI, confidence interval. VISN, Veterans Integrated Service Networks.

Characteristics by treatment group are shown in **Table 1**. Median ages were 61.7 years in the no treatment group and 67, 71.8, and 72.4 years in the nirmatrelvir-ritonavir, molnupiravir, and monoclonal antibody groups, respectively. There were 24,495 (10.2%) persons of Hispanic ethnicity in the no treatment group and 2,437 (9.1%), 694 (7.7%), and 336 (7.0%) in the nirmatrelvir-ritonavir, molnupiravir, and monoclonal antibody groups, respectively. There were 165,499 (72.4%) persons of White race in the no treatment group and 18,615 (72%), 6,742 (76.3%), 3,747 (79.8%) in the nirmatrelvir-ritonavir, molnupiravir, and monoclonal antibody groups, respectively. There were 51,537 (22.5%) persons of Black race in the no treatment group and 6,045 (23.4%), 1,773 (20.1%), 786 (16.7%) in the nirmatrelvir-ritonavir, molnupiravir, and monoclonal antibody groups, respectively.

The proportion of veterans who completed primary or booster vaccination was highest in the molnupiravir group [7,707 (85.6%)], followed by nirmatrelvir-ritonavir [12,010 (82.5%)], monoclonal antibody [3,547 (74.1%)], and no treatment [17,949 (71.7%)] groups. The proportion of veterans with a CCI ≥4 was highest in the monoclonal antibody group [1,733 (36.3%], followed by molnupiravir [2,737 (30.4%)], nirmatrelvir-ritonavir [3,928 (14.7%)], and no treatment [32,892 (13.7%)] groups. Monoclonal antibody and molnupiravir groups had the highest prevalence of cardiovascular disease [2,601 (54.4%) monoclonal antibody, 4,819 (53.5%) molnupiravir, 9,085 (34.1%) nirmatrelvir-ritonavir, 71,345 (29.8) no treatment]; chronic kidney disease [2,302 (48.1%) monoclonal antibody, 4,059 (45.1%) molnupiravir, 9,346 (35.0%) nirmatrelvir-ritonavir, 70,607 (29.5%) no treatment]; and immunocompromised individuals [1,204 (25.2%) monoclonal antibody, 1,210 (13.4%) molnupiravir, 2,653 (9.9%) nirmatrelvir-ritonavir, 17,881 (7.5) no treatment].

### Temporal Trends in COVID-19 Treatments

The proportion of test-positive patients who received any treatment increased from 3.2% (3,285 of 102,343) in January 2022 to a peak of 23.9% (5,180 of 21,688) in August 2022 and was 20.8% (2,194 of 10,551) by January 2023 (**Figure 2**). The proportion of test-positive patients who received nirmatrelvir-ritonavir increased from 1.0% (1,074 of 102,343) in January to 16.8% (3,649 of 21,688) in August 2022 and was 14.5% (1,531 of 10,551) by January 2023 (**Figure 2**). Reductions were most notable in the oldest (≥75 years) age group and among White and Hispanic veterans (**Supplemental Figure 2**). The proportion of test-positive patients who received molnupiravir increased from 0.7% (676 of 102,343) in January to 5.4% (1,167 of 21,688) in August and was 6.3% (663 of 10,551) by January 2023. The proportion treated with monoclonals was highest in February [3.5% (706 of 20,450)], gradually declined to 0.8% (643 of 12,296) by November, and were no longer prescribed as of December 2022.

### Regional Patterns in COVID-19 Treatments

There was substantial variation by VISN in the proportion of test-positive patients who received nirmatrelvir-ritonavir and molnupiravir, which increased notably in April 2022 and persisted through the end of the study (**Figure 3**). By January 2023, the range in proportions was 5.9 to 21.4% in the nirmatrelvir-ritonavir group and 2.1 to 11.1% in the molnupiravir group. The ratio of veterans receiving nirmatrelvir-ritonavir relative to molnupiravir during January 2023 varied from 1.0 to 6.8 across the different VISNs, suggesting that more nirmatrelvir-ritonavir was prescribed at the VISN level. There was also large variability in the proportions of veterans prescribed nirmatrelvir-ritonavir and molnupiravir across VA facilities (**Supplemental Figure 2**). Of 130 facilities which prescribed either oral antiviral, 12 (9%) prescribed more molnupiravir compared with nirmatrelvir-ritonavir.

**Figure 3.**
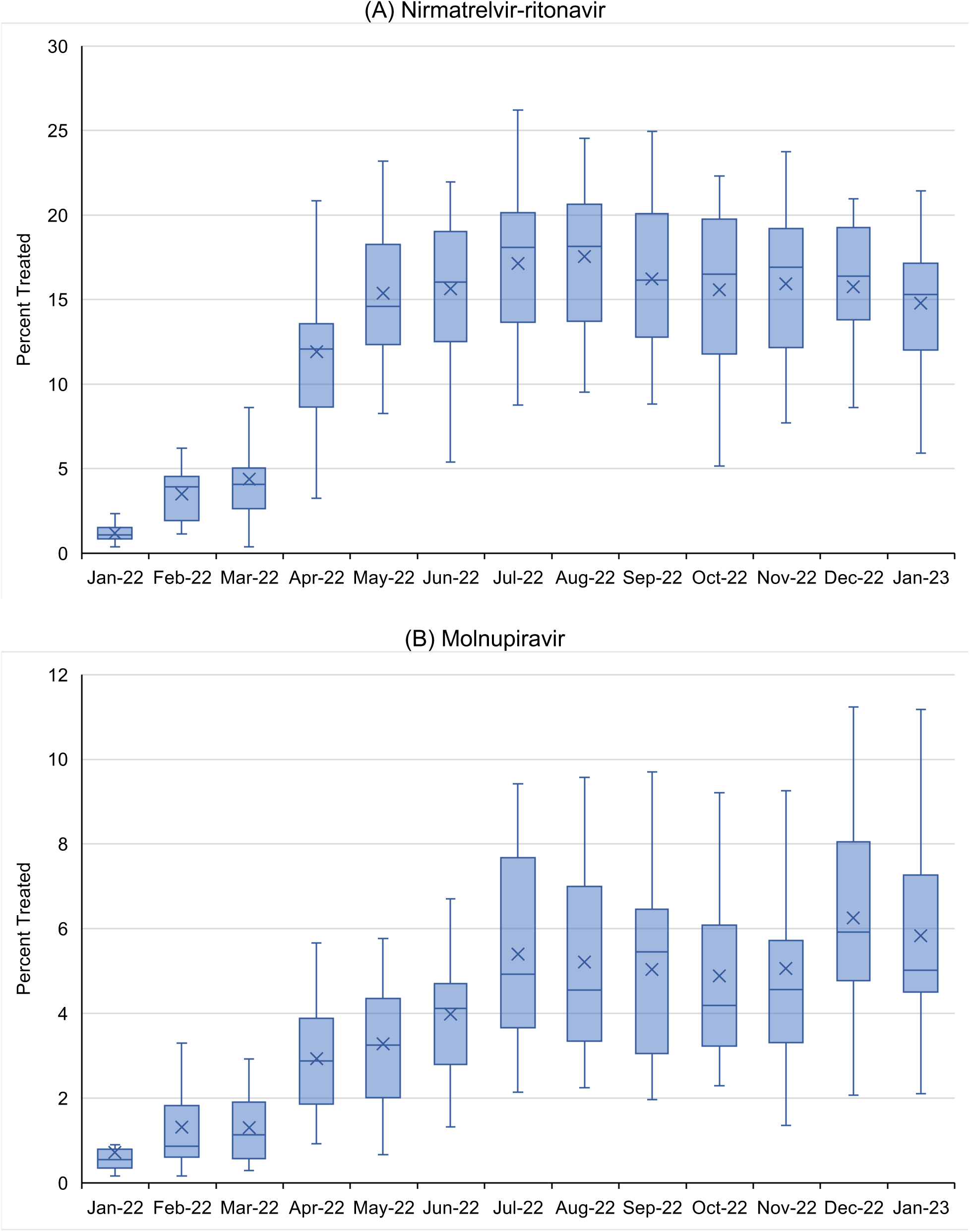
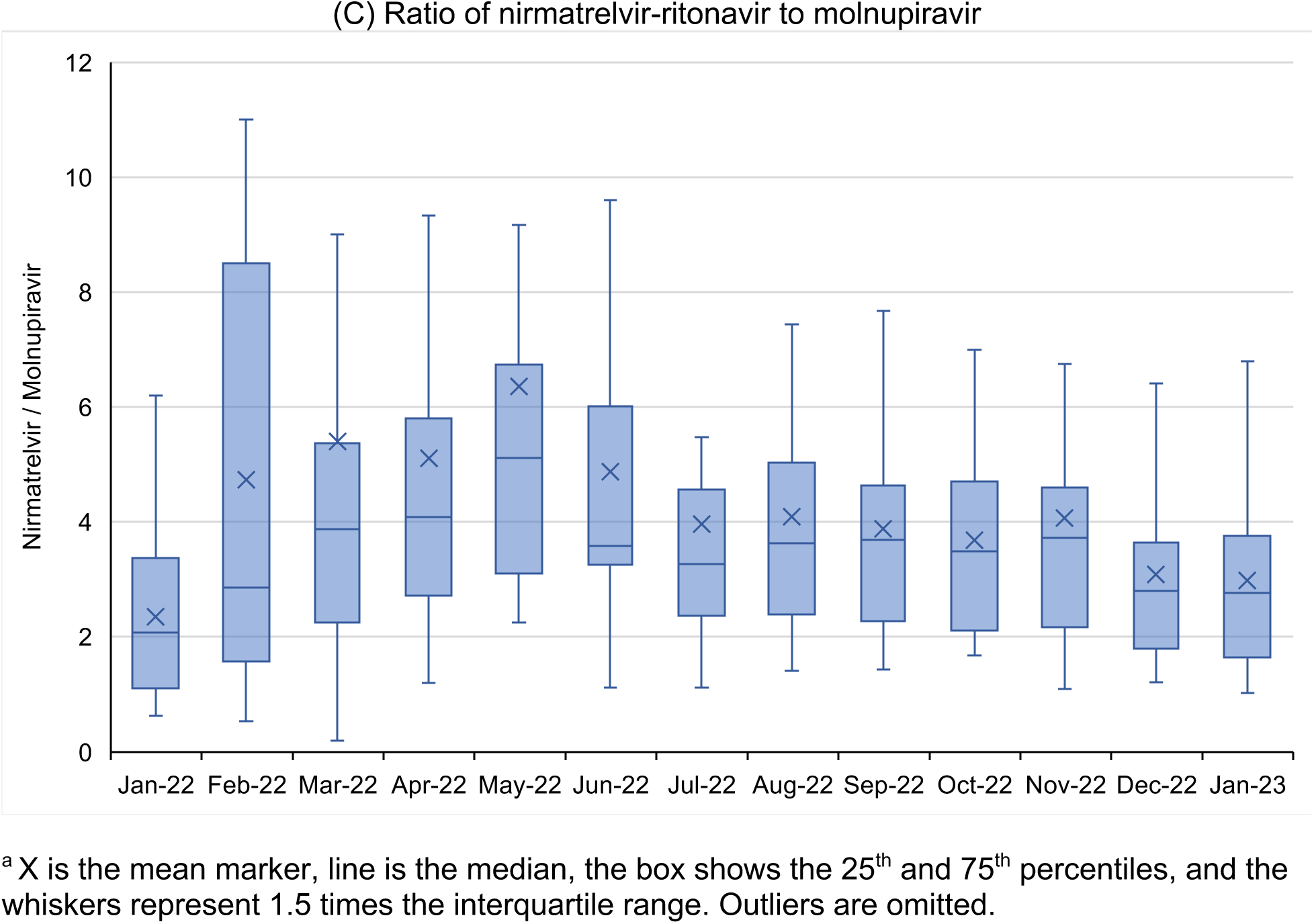
Distribution of COVID-19 pharmacotherapies across the VA’s 18 Veterans Integrated Services Networks (VISNs) by month^a^

### Factors Associated with COVID-19 Treatment

Between April 2022 and January 2023, factors independently associated with higher likelihood of COVID-19 treatment were older age (aOR 1.18, 95% CI 1.14-1.22 for 65-74 versus 50-64 years; aOR 1.19, 95% CI 1.15-1.23 for ≥75 versus 50-64 years), Black vs. White race (aOR 1.06, 95% CI 1.02-1.09) and Hispanic ethnicity (aOR 1.06, 95% CI 1.01-1.11), higher CCI score (aOR 1.39, 95% CI 1.33-1.45 for 4-5 versus 0; aOR 1.52, 95% CI 1.44-1.59 for ≥6 versus 0), and receipt of vaccination vs. no vaccination (aOR 1.25, 95% CI 1.19-1.30 primary vaccination; aOR 1.47, 95% CI 1.42-1.53 booster vaccination). Factors independently associated with lower likelihood of treatment included rural vs. urban residence (aOR 0.84, 95% CI 0.82-0.87), current smoking (aOR 0.84, 95% CI 0.81-0.88), alcohol use disorder (aOR 0.90, 95% CI 0.87-0.92), and substance use disorder (aOR 0.92, 95% CI 0.86-0.98). Compared to veterans in VISN 1, those enrolled in all other VISNs except VISN 9 were less likely to receive treatment.

## Discussion

Among the 285,710 non-hospitalized U.S. veterans who tested positive for SARS-CoV-2 between January 2022 and January 2023, the proportion receiving any outpatient pharmacotherapy increased substantially from 3.2% to 23.9% between January and August 2022 and decreased to 20.8% by January 2023. Although nirmatrelvir-ritonavir remains the most prescribed treatment, the ratio of nirmatrelvir-ritonavir to molnupiravir prescribing has steady decreased from May 2022 to January 2023. There were notable regional differences by VISN in the relative use of different pharmacotherapies. Older Veterans with a higher burden of underlying conditions as well as persons of Black race and Hispanic ethnicity were more likely to receive treatment, whereas unvaccinated veterans and those living in rural areas were less likely to receive treatment.

Several factors may have contributed to the overall decline in the proportion of veterans receiving treatment after August 2022. The absence of available CMS Medicare or VA Community Care data after September 2022 may explain only a part of the decline,since these data only contributed an additional 1% to the monthly treatment ratebetween January and September 2022. A relative increase in asymptomatic infections or milder disease could have occurred in the setting of improved COVID-19 vaccination or previous infection.^21^ Furthermore, overall reductions in COVID-19-related hospitalizations and deaths as well as relaxation of restrictions implemented during the pandemic may have influenced risk perception and care-seeking behavior.^22^ Changes in infrastructure supporting COVID-19 care could also impact prescribing. Although trends in early uptake of COVID-19 pharmacotherapies within VHA mirrored other settings, a more recent decline has not been widely reported in non-veteran populations.^9,23^

The overall small decline in treatment was largely driven by reduced nirmatrelvir-ritonavir dispensing, which remained the most prescribed pharmacotherapy. Possible reasons for this observation include provider and patient concerns about COVID-19 rebound following completion of nirmatrelvir-ritonavir as well as challenges in identifying and managing drug-drug interactions.^4^ In contrast, dispensing of molnupiravir increased during the study period, most notably after FDA authorization for monoclonal antibodies was withdrawn in the setting of reduced susceptibility of circulating Omicron variants. Molnupiravir recipients, similar to patients who had received monoclonal antibodies, were older and had a higher burden of underlying conditions compared with other groups and therefore may not have been eligible to receive nirmatrelvir-ritonavir due to renal or drug-drug contraindications.^24^

Use of COVID-19 pharmacotherapies varied greatly across VISNs and VA facilities and was not limited by distribution of drug supplies to VHA pharmacies. Despite nirmatrelvir-ritonavir being preferentially recommended over molnupiravir and having more robust evidence supporting its clinical effectiveness, a number of facilities still prescribed more molnupiravir than nirmatrelvir-ritonavir.^1,3,5,6,23,25^ Altogether, these differences may be attributed to regional and local variation in policy, infrastructure, education, and provider preferences. For example, higher likelihood of treatment in the VA New England Healthcare System (VISN 1) compared with nearly all other VISNs may have been a result of the regional Test-To-Treat pilot program.

Our previous work showed that Black and Hispanic veterans who tested positive for SARS-CoV-2 during January and February 2022 were less likely to receive outpatient COVID-19 treatment.^8^ However, over an expanded period of time, Black and Hispanic veterans are now slightly more likely to receive treatment. This suggests important progress in improving outreach to these minority racial and ethnic groups. Moreover, compared with non-veteran populations, racial and ethnic disparities in COVID-19-related care within VHA are often less pronounced.^10^

The association of older age and underlying conditions with receipt of COVID-19 treatment is consistent with evidence that these groups are at higher risk for severe COVID-19 outcomes.^26,27^ A treatment pattern consistent with higher risk was not observed among unvaccinated veterans, who were less likely to receive any pharmacotherapy. This may reflect differences in patient behaviors, with veterans who seek vaccination also more likely to pursue treatment for COVID-19. In contrast to our earlier findings, rural veterans were less likely to receive any pharmacotherapy, which may reflect challenges in meeting a higher volume of oral antiviral treatment demand over time.^8^ It is also possible that rural veterans were more likely to seek care outside of VHA, and although we included data from VA Community Care and CMS-Medicare claims, we may still have underascertained prescribing in this group. Persons with alcohol and substance use disorders were also less likely to receive treatment, indicating a need for targeted outreach to these vulnerable groups.

This study has several limitations. First, as previously described, we could not fully ascertain whether veterans were symptomatic at the time of testing positive and truly eligible for treatment under EUA criteria.^8^ Trends in prescribing may have been impacted by changes in the relative proportion of asymptomatic infections over time. Second, although we integrated multiple databases, including VHA CDW, CMS-Medicare, and VA Community Care claims to determine receipt of COVID-19 pharmacotherapies, we may not have accounted for all treatments provided outside of VHA, particularly among veterans younger than 65 years not enrolled in Medicare. In addition, data from CMS-Medicare and VA Community Care were only available through September 2022. However, the relative contributions to ascertainment of COVID-19 treatments between January and September 2022 were small and fairly stable. Third, we did not capture positive SARS-CoV-2 laboratory tests performed outside VHA nor results of self-testing not documented in VHA clinical records. Given our study eligibility criteria, this would have impacted measurement of additional infections as well as COVID-19 treatments. Fourth, we were not able to enumerate the number of veterans who were offered but declined treatment. Finally, given differences in veteran demographic and clinical characteristics as compared with the general US population as well as differences in care delivery between VHA and non-VHA systems, findings from this study may not be generalizable to other groups.

## Conclusions

In this nationwide study of US veterans in VHA care who tested positive for SARS-CoV-2 between January 2022 and January 2023, prescription of outpatient COVID-19 pharmacotherapies steadily increased until August 2022 and declined slightly thereafter. This occurred in the setting of a relative increase in molnupiravir use, a reduction in nirmatrelvir-ritonavir use, and cessation of monoclonal antibody administration following removal of FDA authorization. The likelihood of receiving treatment was influenced by demographic, clinical, and geographic differences in systems of care. These results highlight important progress in reaching certain groups, including Black and Hispanic veterans. They also demonstrate the need for continued support of infrastructure and education to facilitate treatment for the highest-risk individuals who may progress to severe COVID-19. Future research should focus on evaluating the relative benefits and risks of available outpatient COVID-19 pharmacotherapies across different patient populations.

## Data Availability

All data produced in the present work are contained in the manuscript.

## Disclaimer

The contents of this article do not represent the views of the US Department of Veterans Affairs or the US Government.

## Additional Contributions

We thank the Biomedical Advanced Research and Development Authority (BARDA, IAA#: AAI21050) and US Food and Drug Administration (FDA, IAA#: 75F40121S30013) for their support.

## Supplemental Material

**Supplemental Table 1.** Distribution of COVID-19 pharmacotherapies by month and data source. Most of the treatments were identified through Veterans Affairs Corporate Data Warehouse (VA CDW). A small fraction of the treatments (7-153 courses of nirmatrelvir-ritonavir, 0-45 of courses molnupiravir, and 19-158 courses of monoclonals depending on the month) were identified through Centers for Medicare & Medicaid Services (CMS). The VA Community Care program contributed a few additional monoclonal antibodies.

|  |  | Jan-2022 | Feb | Mar | Apr | May | Jun | Jul | Aug | Sept | Oct | Nov | Dec | Jan-2023 |
| --- | --- | --- | --- | --- | --- | --- | --- | --- | --- | --- | --- | --- | --- | --- |
| SARS-CoV-2 Infection |  | 102,343 | 20,450 | 5,724 | 6,249 | 16,089 | 21,804 | 26,985 | 21,688 | 14,088 | 9,727 | 12,296 | 17,716 | 10,551 |
| Prescriptions In VA CDW | Nirmatrelvir-ritonavir | 1062 | 654 | 244 | 812 | 2359 | 3111 | 4210 | 3507 | 2092 | 1524 | 2017 | 2822 | 1531 |
|  | Molnupiravir | 652 | 280 | 74 | 192 | 550 | 860 | 1398 | 1122 | 709 | 480 | 643 | 1153 | 663 |
|  | Monoclonals | 1200 | 556 | 141 | 89 | 275 | 350 | 470 | 284 | 226 | 148 | 84 | 0 | 0 |
| Additional<sup>a</sup> Prescriptions in CMS<sup>b</sup> | Nirmatrelvir-ritonavir or Molnupiravir | 36 | 23 | <11* | 24 | 110 | 153 | 197 | 186 | 161 | - | - | - | - |
|  | Monoclonals | 158 | 74 | 22 | 19 | 24 | 35 | 34 | 31 | 23 | - | - | - | - |
| Additional<sup>a</sup> Prescriptions In Community Care<sup>b</sup> | Nirmatrelvir-ritonavir or Molnupiravir | 0 | 0 | 0 | 0 | 0 | 0 | 0 | 0 | 0 | - | - | - | - |
|  | Monoclonals | 177 | 76 | 22 | 10 | 38 | 47 | 67 | 49 | 30 | - | - | - | - |

<sup>a</sup> These include prescriptions not already captured in VA CDW.
<sup>b</sup> Prescription data from CMS and Community care are only available through September 2022.
\* Data from CMS were combined and masked in accordance with CMS cell size suppression policy (https://resdac.org/articles/cms-cell-size-suppression-policy).

**Supplemental Figure 1.**
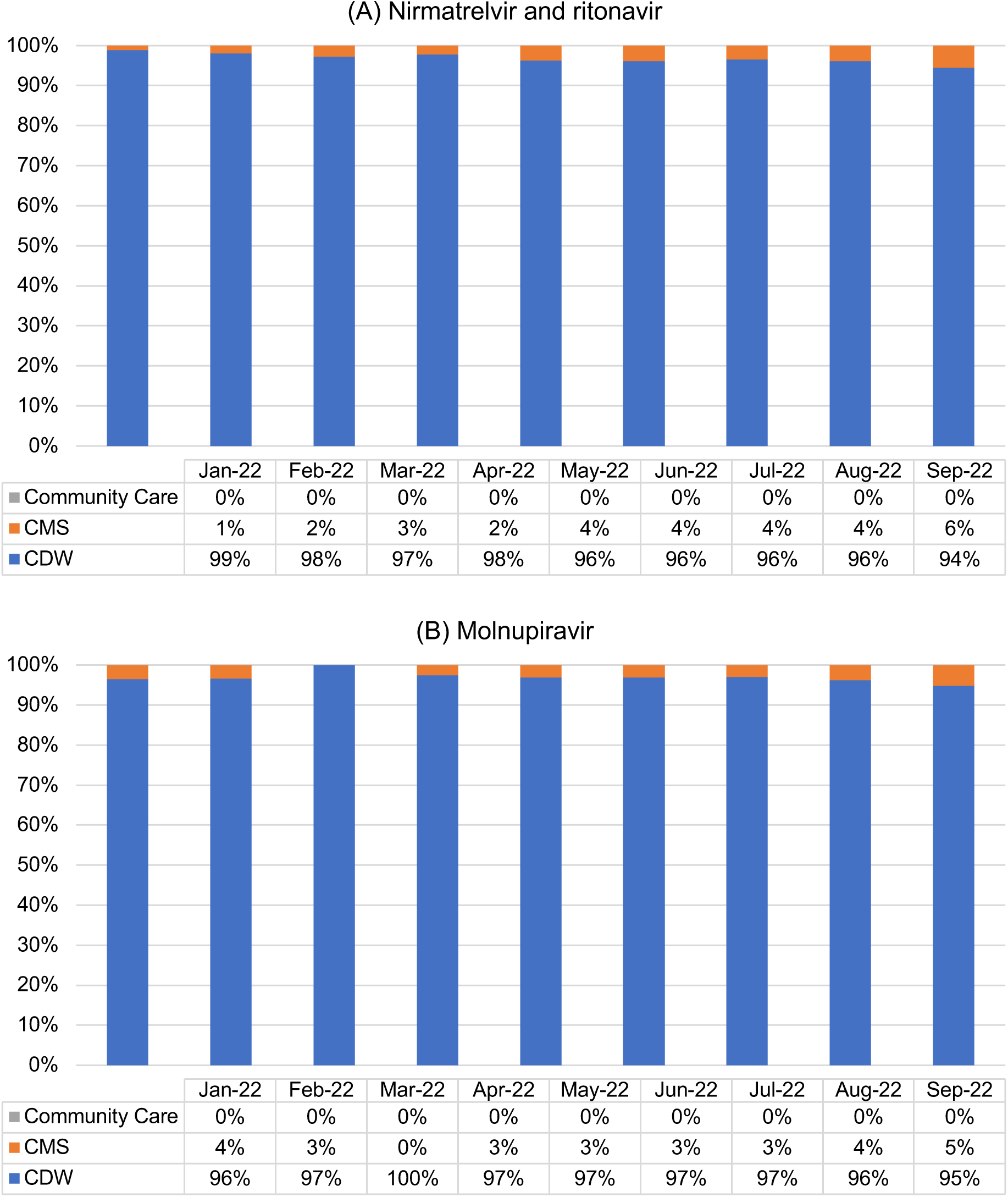
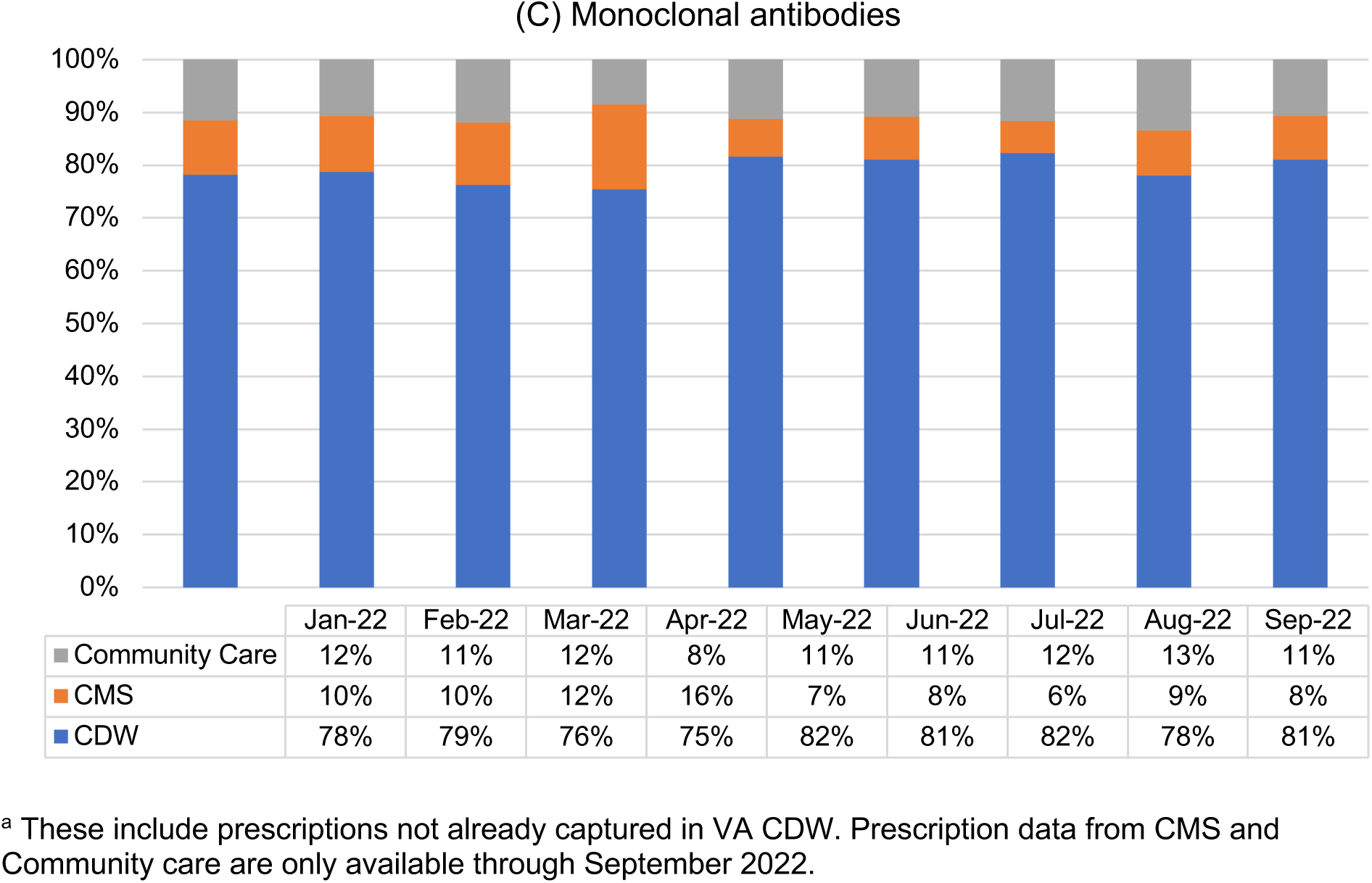
Distribution of COVID-19 pharmacotherapies by data source^a^

**Supplemental Figure 2.**
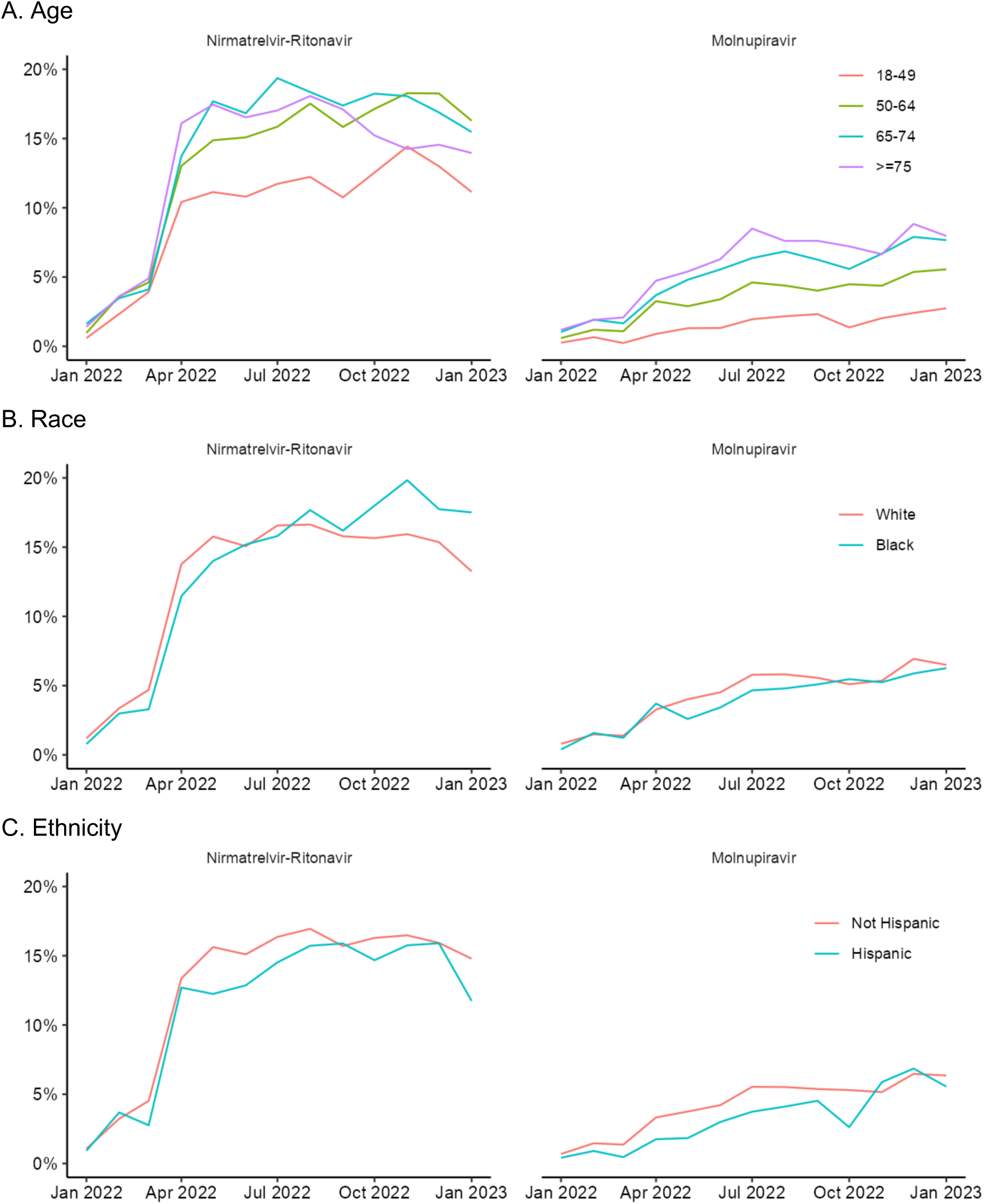
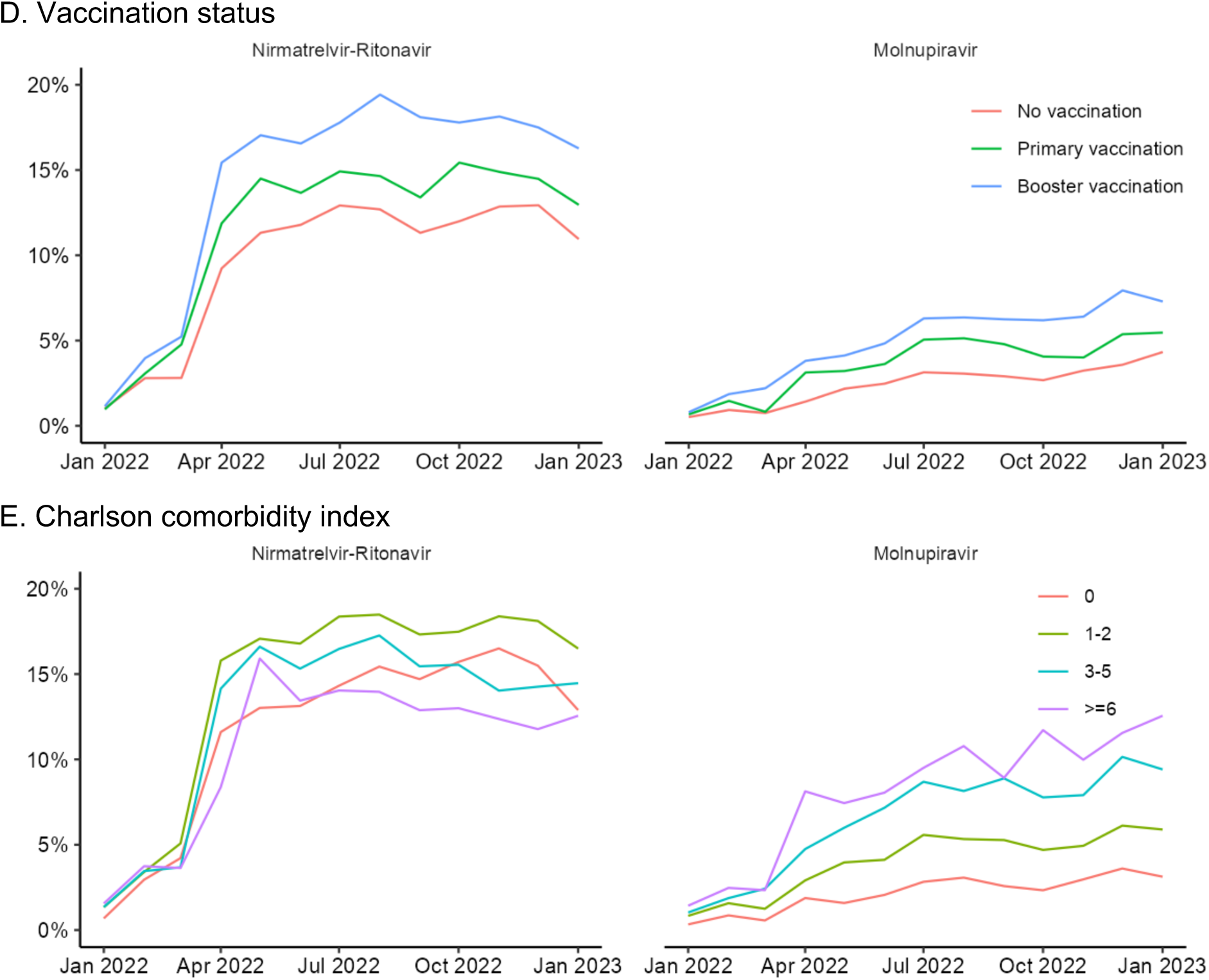
Monthly receipt of nirmatrelvir-ritonavir and molnupiravir by demographic and clinical characteristics

**Supplemental Figure 3.**
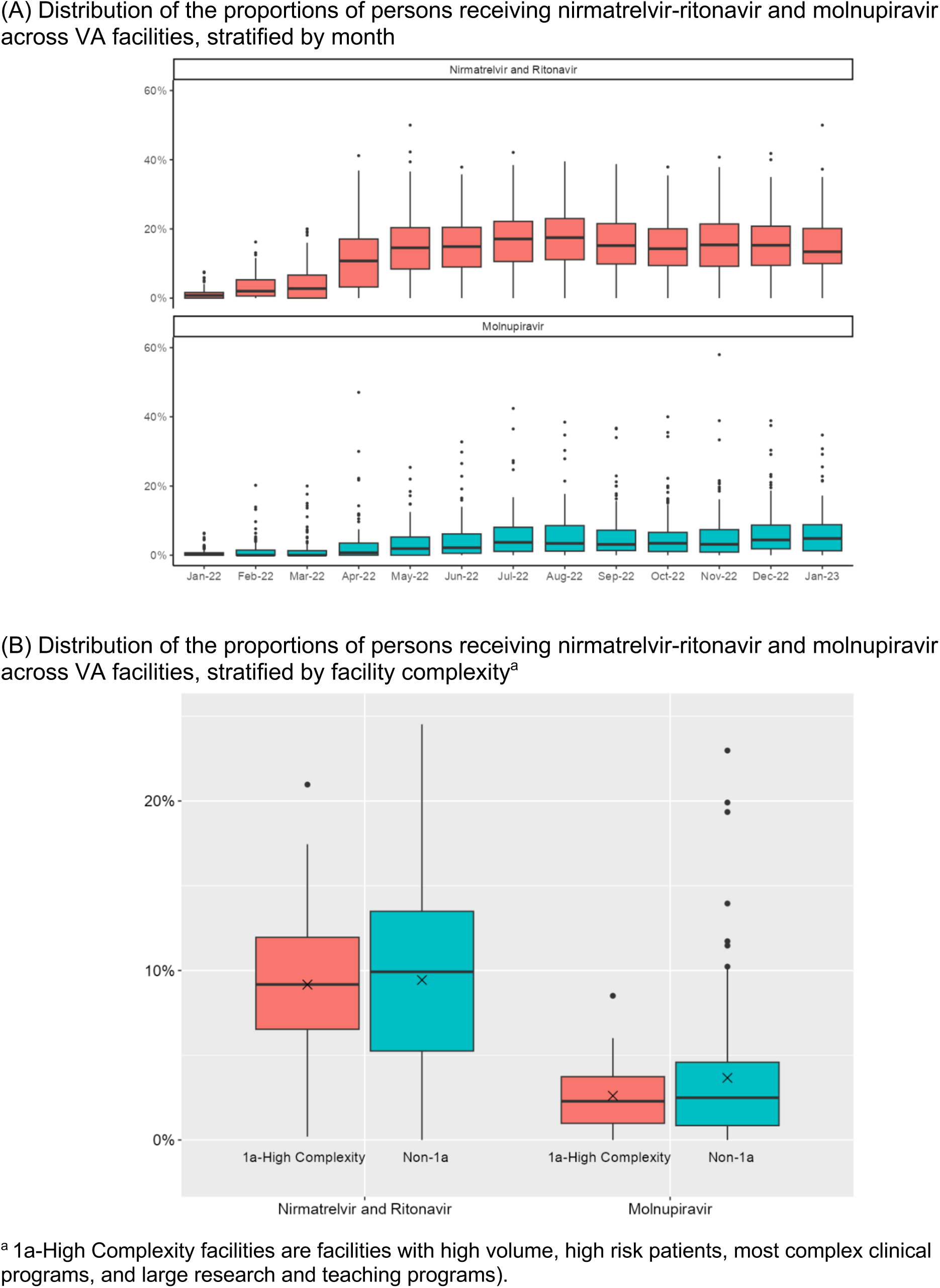
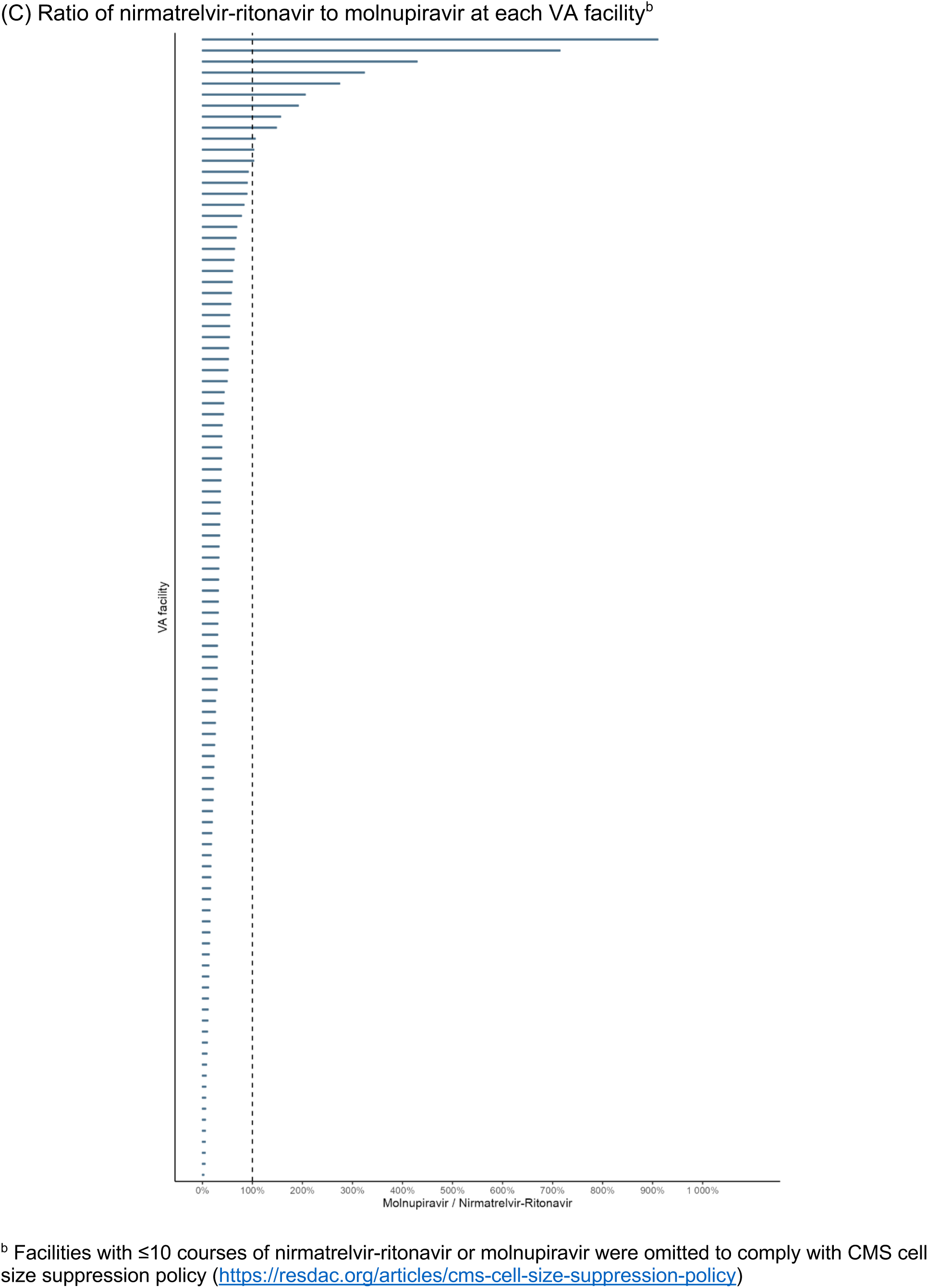
Distribution of COVID-19 pharmacotherapies by facility

## References

1. National Institutes of Health. Therapeutic Management of Nonhospitalized Adults With COVID-19. https://www.covid19treatmentguidelines.nih.gov/management/clinical-management-of-adults/nonhospitalized-adults--therapeutic-management/ Published 2022. Accessed March 21, 2023.

2. U.S. Food and Drug Administration. Coronavirus (COVID-19) | Drugs. https://www.fda.gov/drugs/emergency-preparedness-drugs/coronavirus-covid-19-drugs. Published 2023. Accessed March 21, 2023.

3. Hammond J, Leister-Tebbe H, Gardner A, et al. Oral Nirmatrelvir for High-Risk, Nonhospitalized Adults with Covid-19. N Engl J Med. 2022;386(15):1397–1408.

4. U.S. Food and Drug Administration. FDA Briefing Document: NDA #217188. https://www.fda.gov/media/166197/download. Published 2023. Accessed March 21, 2023.

5. Butler CC, Hobbs FDR, Gbinigie OA, et al. Molnupiravir plus usual care versus usual care alone as early treatment for adults with COVID-19 at increased risk of adverse outcomes (PANORAMIC): an open-label, platform-adaptive randomised controlled trial. Lancet. 2023;401(10373):281–293.

6. Jayk Bernal A, Gomes da Silva MM, Musungaie DB, et al. Molnupiravir for Oral Treatment of Covid-19 in Nonhospitalized Patients. N Engl J Med. 2022;386(6):509–520.

7. Gottlieb RL, Vaca CE, Paredes R, et al. Early Remdesivir to Prevent Progression to Severe Covid-19 in Outpatients. N Engl J Med. 2022;386(4):305–315.

8. Bajema KL, Wang XQ, Hynes DM, et al. Early Adoption of Anti–SARS-CoV-2 Pharmacotherapies Among US Veterans With Mild to Moderate COVID-19, January and February 2022. JAMA Network Open. 2023;5(11).

9. Gold JAW, Kelleher J, Magid J, et al. Dispensing of Oral Antiviral Drugs for Treatment of COVID-19 by Zip Code-Level Social Vulnerability - United States, December 23, 2021-May 21, 2022. MMWR Morb Mortal Wkly Rep. 2022;71(25):825–829.

10. Boehmer TK, Koumans EH, Skillen EL, et al. Racial and Ethnic Disparities in Outpatient Treatment of COVID-19 - United States, January-July 2022. MMWR Morb Mortal Wkly Rep. 2022;71(43):1359–1365.

11. Administration for Strategic Preparedness & Response (ASPR). COVID-19 Therapeutics Thresholds, Orders, and Replenishment by Jurisdiction. U.S. Department of Health and Human Services,. https://aspr.hhs.gov/COVID-19/Therapeutics/orders/Pages/default.aspx. Published 2023. Accessed March 21, 2023.

12. Veterans Health Administration. https://www.va.gov/health/. Published 2022. Accessed March 13, 2023.

13. Bajema KL, Wang XQ, Hynes DM, et al. Early Adoption of Anti-SARS-CoV-2 Pharmacotherapies Among US Veterans With Mild to Moderate COVID-19, January and February 2022. JAMA Netw Open. 2022;5(11):e2241434.

14. U.S. Department of Veterans Affairs. COVID-19:Shared Data Resource. https://vhacdwdwhweb100.vha.med.va.gov/phenotype/index.php/COVID-19:Shared_Data_Resource. Published 2023. Accessed March 21, 2023.

15. U.S. Department of Veterans Affairs. New Tool Helps VA Track and Analyze COVID-19 Data on the Ground. https://www.oit.va.gov/news/article/?read=new-tool-helps-va-track-and-analyze-covid-19-data. Published 2020. Accessed June 3, 2022.

16. Veterans Integrated Services Networks (VISNs). Veterans Health Administration. https://www.va.gov/HEALTH/visns.asp. Accessed March 13, 2023.

17. U.S. Department of Agriculture. Rural-Urban Commuting Area Codes. https://www.ers.usda.gov/data-products/rural-urban-commuting-area-codes/. Published 2022. Accessed May 10, 2022.

18. Bajema KL, Berry K, Streja E, et al. Effectiveness of COVID-19 treatment with nirmatrelvir-ritonavir or molnupiravir among U.S. Veterans: target trial emulation studies with one-month and six-month outcomes. medRxiv. 2022:2022.2012.2005.22283134.

19. Administration for Strategic Preparedness & Response (ASPR). ASPR pauses allocation of bamlanivimab and etesevimab together, etesevimab alone, and REGEN-COV. https://www.phe.gov/emergency/events/COVID19/therapeutics/update-23Dec2021/Pages/default.aspx. Published 2022. Accessed February 23, 2022.

20. Emergency Use Authorization. U.S. Food and Drug Administration. https://www.fda.gov/emergency-preparedness-and-response/mcm-legal-regulatory-and-policy-framework/emergency-use-authorization#coviddrugs. Accessed March 13, 2023.

21. Bajema KL, Rowneki M, Berry K, et al. Rates of and Factors Associated With Primary and Booster COVID-19 Vaccine Receipt by US Veterans, December 2020 to June 2022. JAMA Netw Open. 2023;6(2):e2254387.

22. Centers for Disease Control and Prevention. COVID Data Tracker. https://covid.cdc.gov/covid-data-tracker/?msclkid=75ea855fd09011eca5e2f76fc9ead3e6#datatracker-home. Published 2022. Accessed March 1, 2022.

23. Lewnard JA, McLaughlin JM, Malden D, et al. Effectiveness of nirmatrelvir-ritonavir against hospital admission or death: a cohort study in a large US healthcare system. medRxiv. 2023.

24. U.S. Food and Drug Administration. Fact sheet for health care providers: Emergency use authorization for Paxlovid. https://www.fda.gov/media/155050/download. Published 2023. Accessed April 2, 2023.

25. Dryden-Peterson S, Kim A, Kim AY, et al. Nirmatrelvir Plus Ritonavir for Early COVID-19 in a Large U.S. Health System : A Population-Based Cohort Study. Ann Intern Med. 2023;176(1):77–84.

26. Centers for Disease Control and Prevention. Risk for COVID-19 Infection, Hospitalization, and Death By Age Group. https://www.cdc.gov/coronavirus/2019-ncov/covid-data/investigations-discovery/hospitalization-death-by-age.html#footnote04. Published 2022. Accessed 11 November, 2022.

27. Centers for Disease Control and Prevention. Underlying Medical Conditions Associated with Higher Risk for Severe COVID-19: Information for Healthcare Professionals. https://www.cdc.gov/coronavirus/2019-ncov/hcp/clinical-care/underlyingconditions.html?msclkid=1e40c3e2d09711ec8b9ea081710e6bc2. Published 2022. Accessed May 10, 2022.

